# Diet order affects energy balance in randomized crossover feeding studies that vary in macronutrients but not ultra-processing

**DOI:** 10.1101/2023.10.03.23296501

**Authors:** Christina M. Sciarrillo, Juen Guo, Aaron Hengist, Valerie L. Darcey, Kevin D. Hall

**Author notes:** Corresponding author: Kevin D. Hall.

## Abstract

**BACKGROUND:** Crossover studies can induce order effects, especially when they lack a wash-out period.

**OBJECTIVE:** To explore diet order effects on energy balance and food intake between randomized diet order groups in two inpatient crossover studies originally designed to compare within-subject differences in *ad libitum* energy intake between either minimally processed low carbohydrate (LC) versus low fat (LF) diets or macronutrient-matched diets composed of mostly minimally processed food (MPF) or ultra-processed food (UPF).

**METHODS:** Diet order group comparisons of changes in body weight, body composition, and differences in energy expenditure, and food intake were assessed over four weeks in 20 adults randomized to either the LC followed immediately by the LF diet (LC→LF) or the opposite order (LF→LC) as well as 20 adults randomized to either the MPF followed by UPF (MPF→UPF) diets or the opposite order (UPF→MPF).

**RESULTS:** Subjects randomized to LC→LF lost 2.9 ± 1.1 kg more body weight (*p* < 0.001) and 1.5 ± 0.6 kg more body fat (*p* = 0.03) than the LF→LC group likely because the LC→LF group consumed 922 ± 304 kcal/d less than the LFàLC group (*p* = 0.0024). Reduced energy intake in LC→LF *vs* LFàLC was driven by the last two weeks (−1610 ± 306 kcal/d; *p*<0.00001) perhaps due to carryover effects of gut adaptations over the first two weeks arising from large differences in the mass of food (1295 ± 209 g/d; *p*<0.00001) and fiber intake (58 ± 5 g/d; *p*<0.00001). There were no diet order effects on *ad libitum* energy intake, body weight, or body composition change between UPF→MPF versus MPF→UPF groups.

**CONCLUSIONS:** Diet order influences daily *ad libitum* energy intake, body weight change, and fat change within the context of a 4-week crossover inpatient diet study varying in macronutrients, but not varying in extent and purpose of processing.

**Funding sources:** Intramural Research Program of the National Institute of Diabetes and Digestive and Kidney Diseases, National Institutes of Health

**Clinical Trial Registration:** NCT03407053 and NCT03878108

## INTRODUCTION

Crossover study designs in nutrition research are advantageous because each participant receives all interventions and serves as their own control, which increases statistical power [1]. Such study designs are especially important for inpatient controlled feeding trials, which are limited in size and duration for practical reasons but are necessary to investigate the effects of diets *per se* rather than the effects of diet advice [2]. However, crossover studies are vulnerable to order effects, whereby earlier interventions can carry over to affect subsequent outcomes, especially when washout periods are insufficient to return participants to their baseline state [3].

In our experience, washout periods increase the risk that participants withdraw from the study before completing all interventions and our most recent inpatient crossover studies therefore did not have washout periods [4, 5]. The primary outcomes of those studies were to determine within subject differences in mean daily *ad libitum* energy intake between minimally processed low carbohydrate (LC) versus low fat (LF) diets [5] or between diets with similar macronutrient composition but differing in the content of ultra-processed food (UPF) versus minimally processed food (MPF) [4]. While neither study exhibited significant within-subject diet order effects on the primary outcomes [4, 5], we did not previously evaluate the differences between groups randomized to the different diet orders. Thus, the present study is a secondary analysis to explore the effects of diet order between groups randomized to the LC followed by LF diet (LC → LF) versus the reverse order (LF → LC) as well as the groups randomized to the UPF followed by MPF diet (UPF → MPF) versus the reverse order (MPF → UPF).

## METHODS

### Participants

The study protocols were approved by the Institutional Review Board of the National Institute of Diabetes & Digestive & Kidney Diseases (NCT03878108; NCT03407053). Detailed methods have been published previously and elsewhere [4, 5] but will be discussed briefly below. All participants provided written consent to participate in the study after being fully informed of the risks of the study. All participants were healthy adults, age 18-50 years, male and female, and weight stable (< ± 5% over past 6 months).

### Research setting

All participants were admitted as inpatients to the Metabolic Clinical Research Unit at the NIH Clinical Center between the years 2018 and 2020. Participants resided in individual rooms. Participants could have visitors meet with them in a common area with supervision by research team personnel to avoid exposure to off-study foods or beverages. The present secondary analyses design is depicted in **Figure 1**. For the protocol providing a LC vs. LF diet for 2 weeks each in random order, participants were randomly assigned to one of two sequences: 1) LC diet first for 14 days followed by the LF diet for 14 days (LC → LF); 2) LF diet first for 14 days followed by the LC diet for 14 days (LF → LC). For the protocol providing an ultra-processed (primarily NOVA category 4) vs. minimally processed (primarily NOVA category 1) diet for 2 weeks each in random order, participants were randomly assigned to one of two sequences: 1) ultra-processed diet first for 14 days followed by the minimally processed diet for 14 days (UPF → MPF); 2) minimally processed diet first for 14 days followed by the ultra-processed diet for 14 days (MPF → UPF). Neither study had a wash-out period. Participants resided in individual rooms and consumed all food and beverages in their rooms with the door open, except for days spent in the respiratory chambers. Any visitations were held in a common area with supervision by research team personnel to avoid exposure to off-study foods or beverages.

**Figure 1.**
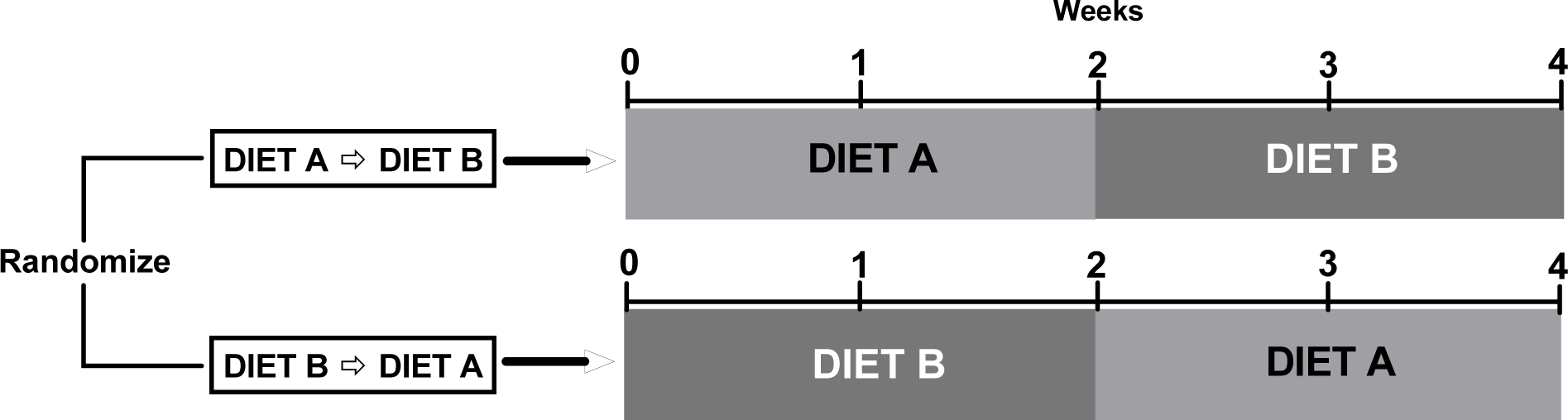
Study Design. The present study is a secondary analysis of two studies [4,5]. The studies were designed to investigate within subject differences in *ad libitum* energy intake between Diet A vs Diet B. In the present secondary analysis, we explored the effects of diet order between the groups randomized to Diet A→ Diet B versus Diet B→Diet A.

### Diets

All meals and snacks for both protocol diets were designed and analyzed using ProNutra software (v.3.4, Viocare) with nutrient values derived from the USDA National Nutrient Database for Standard Reference, Release 26 and the USDA Food and Nutrient Database for Dietary Studies, 4.0. For all diets, 7-day rotating menus were used. Foods and beverages were categorized using the NOVA system regarding the extent and purpose of processing [6] and the glycemic index of foods was calculated relative to 50 g of oral glucose [7]. Both the LC and LF diets were matched for provided energy and non-starchy vegetables. Unlike the LF diet, the LC diet had animal-based products (meat, poultry, fish, eggs, dairy). The LF diet had no animal products and instead included legumes, rice, root vegetables, soy products, corn, lentils, peas, whole grains, bread, and fruit. Both the UPF and MPF diets were matched for provided energy, sugar, carbohydrates, fiber, fat, glycemic load, overall energy density, and sodium. For both protocols, diet-specific snacks and water were provided *ad libitum* during the day in snack boxes located in patient rooms. Meals were presented to patients with instructions to eat as much or as little as desired. All subjects were given 60 minutes to complete their meals. More specific and detailed diet and menu information for both protocols is published elsewhere [4, 5].

After each meal, the remaining food and beverages were identified and weighed by nutrition staff to quantify the amount of each food consumed. Nutrient and energy intake were calculated using ProNutra (v.3.4). This same procedure was also completed for daily water and snacks too.

### Meal tests, Continuous Glucose Monitoring, and Ketone measurements

For the LC and LF diets, after an overnight fast and during the second week of each diet, participants completed a mixed meal test in which they consumed a liquid meal matching the macronutrient content of the current diet and containing 30% of estimated daily calorie requirements. Blood samples were collected at 0, 10, 20, 30, 60, 90, 120, 180, 240, 300, and 360 minutes to quantify postprandial metabolites (c-peptide, free-fatty acids [FFA], glucose, triglycerides, lactate and insulin) and appetite hormones (active glucagon-like peptide 1 [GLP-1], active ghrelin, total ghrelin, glucose-dependent insulinotropic polypeptide [GIP], peptide YY [PYY], leptin). Overnight fasting capillary β-hydroxybutyrate was measured daily during both LC and LF diets using Abbott Precision Xtra blood glucose and ketone monitoring system (Abbott Diabetes Care) in daily finger-prick blood samples obtained from 15 participants. 24-h urinary excretion of β-hydroxybutyrate was measured at the end of each diet period for all 20 participants. Plasma total ketones, β-hydroxybutyrate, acetoacetate, and acetone were measured at baseline and at the end of each diet period for 19 participants. For the UPF and MPF diets, no meal tests (e.g. postprandial appetite hormones or postprandial metabolites), capillary, urine, or plasma β-hydroxybutyrate, were measured during the UPF or MPF diets. For both studies, subjects wore the Dexcom G4 Platinum continuous glucose monitor (CGM) daily.

### Subjective measures of appetite

Subjects completed hunger and satiety assessments on various study days as well as sensory and palatability assessments; namely pleasantness, familiarity, hunger, satisfaction, fullness, and eating capacity. Data were collected using a visual analog scale (VAS) with ratings from 0-100. Detailed information on the collection of these measures is reported elsewhere [4, 5].

### Diet History Questionnaire

The Diet History Questionnaire (DHQ)-III was administered at screening. The DHQ-III is a web-based food frequency questionnaire that queries frequency and typical portion size of 135 foods and beverages and 26 dietary supplements over the past year.

### Body weight and composition

Body weight was measured daily after the first void (Welch Allyn Scale-Tronix 5702; Skaneateles Falls, NY, USA). Body composition measurements were performed at baseline and weekly for the duration of the study using dual-energy X-ray absorptiometry (General Electric Lunar iDXA; Milwaukee, WI, USA).

### Energy Expenditure

For both protocols, subjects resided in respiratory chambers for one day each week. All chamber measurement periods were >23 hours and data were extrapolated to represent 24h periods by assuming that the mean of the measured periods was representative of the 24h period. Energy expenditure was calculated using equations reported elsewhere [4, 5]. Sleeping energy expenditure was determined as the lowest energy expenditure over a continuous 180-min period between the hours of 00:00 and 06:00 [8]. Sedentary energy expenditure includes the thermic effect of food as previously described [9] and physical activity expenditure was the difference between 24h energy expenditure and sedentary energy expenditure.

### Statistical analyses

This secondary analysis was performed using SAS (version 9.4; SAS Institute, Cary, NC, USA). All data are presented as mean ± SE. Data were analyzed by repeated measures mixed model with subject as random effect and diet and diet order were fixed effects (PROC MIXED, SAS). For the scatter plots relating mean daily intake of either energy or carbohydrate to various ketone measurements at the individual subject level, intake data are adjusted for the baseline resting energy expenditure of each subject. Mean plasma concentrations of gut hormones following the LC and LF diets were calculated by dividing total area under the curve (tAUC) by 360 minutes. The conversion factor used for GLP-1 was 1 pmol·L^−1^ = 3.297 pg·mL^−1^. Given the exploratory nature of this secondary analysis, we did not adjust p-values for multiple comparisons.

## RESULTS

Participants randomized to the different diet order groups were similar in mean age (years), BMI (kg/m^2^), fat mass (%), fat mass (kg), respiratory quotient (RQ), resting energy expenditure (REE; kcal/d), or energy intake (kcal/d) between randomized groups for either study (**Table 1**). Whereas there were no differences in habitual healthy eating index (HEI) between UPF → MPF vs. MPF → UPF groups, the LC → LF group had a lower HEI compared to LF → LC group (*p* = 0.02).

**Table 1.** Baseline Participant Characteristics. Data are mean ± SEM. All data are results of unpaired t-tests, except for comparisons of sex between group, where chi-squared test was used. *Data were self - reported and collected using validated diet history questionnaire (DHQ) methods. Abbreviations: **LC** Low-carbohydrate; **LF** Low-fat; **MPF** Minimally processed food; **UPF** Ultra-processed food; **REE** Resting energy expenditure; **CHO** carbohydrate.

|  | LC vs LF diet study |  |  | UPF vs MPF diet study |  |  |
| --- | --- | --- | --- | --- | --- | --- |
|  | LC → LF | LF → LC | *p*-value | MPF → UPF | UPF → MPF | *p*-value |
| **Demographics** |  |  |  |  |  |  |
| Male/female (N) | 5/5 | 6/4 | 0.65 | 4/6 | 6/4 | 0.37 |
| Age (years) | 28 ± 2 | 32 ± 2 | 0.25 | 30 ± 3 | 33 ± 2 | 0.50 |
| BMI (kg/m<sup>2</sup>) | 28.5 ± 2.1 | 26.9 ± 1.7 | 0.57 | 27.9 ± 2.1 | 26.7 ± 2.2 | 0.57 |
| Fat mass (%) | 34.8 ± 9.8 | 30.8 ± 9.7 | 0.37 | 31.5 ± 0.0 | 28.5 ± 0.0 | 0.55 |
| Fat mass (kg) | 28.7 ± 4.0 | 25.0 ± 3.1 | 0.48 | 26.0 ± 4.5 | 23.1 ± 4.2 | 0.64 |
| REE (kcal/d) | 1569 ± 102 | 1563 ± 85 | 0.83 | 1506 ± 95 | 1514 ± 83 | 0.95 |
| Respiratory quotient | 0.803 ± 0.022 | 0.804 ± 0.010 | 0.97 | 0.826 ± 0.017 | 0.841 ± 0.017 | 0.53 |
| **Diet history\*** |  |  |  |  |  |  |
| Energy intake (kcal/d) | 1808 ± 292 | 1757 ± 429 | 0.92 | 1639 ± 272 | 1796 ± 299 | 0.70 |
| Total fat intake (g/d) | 70.9 ± 11.8 | 68.5 ± 19.4 | 0.91 | 66.3 ± 9.2 | 69.7 ± 12.4 | 0.84 |
| Total CHO intake (g/d) | 225.2 ± 38.7 | 213.0 ± 43.8 | 0.84 | 209.9 ± 52.1 | 207.4 ± 31.5 | 0.97 |
| Total protein intake (g/d) | 72.2 ± 12.23 | 76.5 ± 22.9 | 0.87 | 68.6 ± 10.4 | 75.3 ± 13.6 | 0.72 |
| Total added sugar intake (g/d) | 56.9 ± 16.1 | 39.7 ± 6.7 | 0.33 | 8.9 ± 3.4 | 11.7 ± 3.3 | 0.58 |
| Total fiber intake (g/d) | 19.0 ± 3.7 | 22.7 ± 5.9 | 0.60 | 18.8 ± 4.4 | 17.5 ± 4.3 | 0.84 |
| Healthy eating index (0-100) | 61.6 ± 3.3 | 71.8 ± 2.3 | 0.02 | 66.4 ± 3.0 | 64.4 ± 2.2 | 0.59 |

### Comparison between LC→ LF versus LF → LC diet orders

#### Energy Expenditure

Mean 24-hr energy expenditure did not significantly differ between subjects randomized to LC → LF (2099 ± 125 kcal/d) vs. LF → LC (2339 ± 125 kcal/d; *p* = 0.18) (**Figure 2A**). Likewise, sleeping (1388 ± 97 kcal/d for LC → LF vs. 1579 ± 102 kcal/d for LF → LC; *p* = 0.18) and physical activity energy expenditure (380 ± 45 kcal/d for LC → LF vs. 411 ± 48 kcal/d for LF → LC; *p* = 0.64) did not differ by diet order group. Daily METs measured using accelerometry were slightly greater in the LF → LC group (1.52 ± 0.01) compared to LC → LF group (1.49 ± 0.01; *p* = 0.03) (**Figure 2B**).

**Figure 2A-F.**
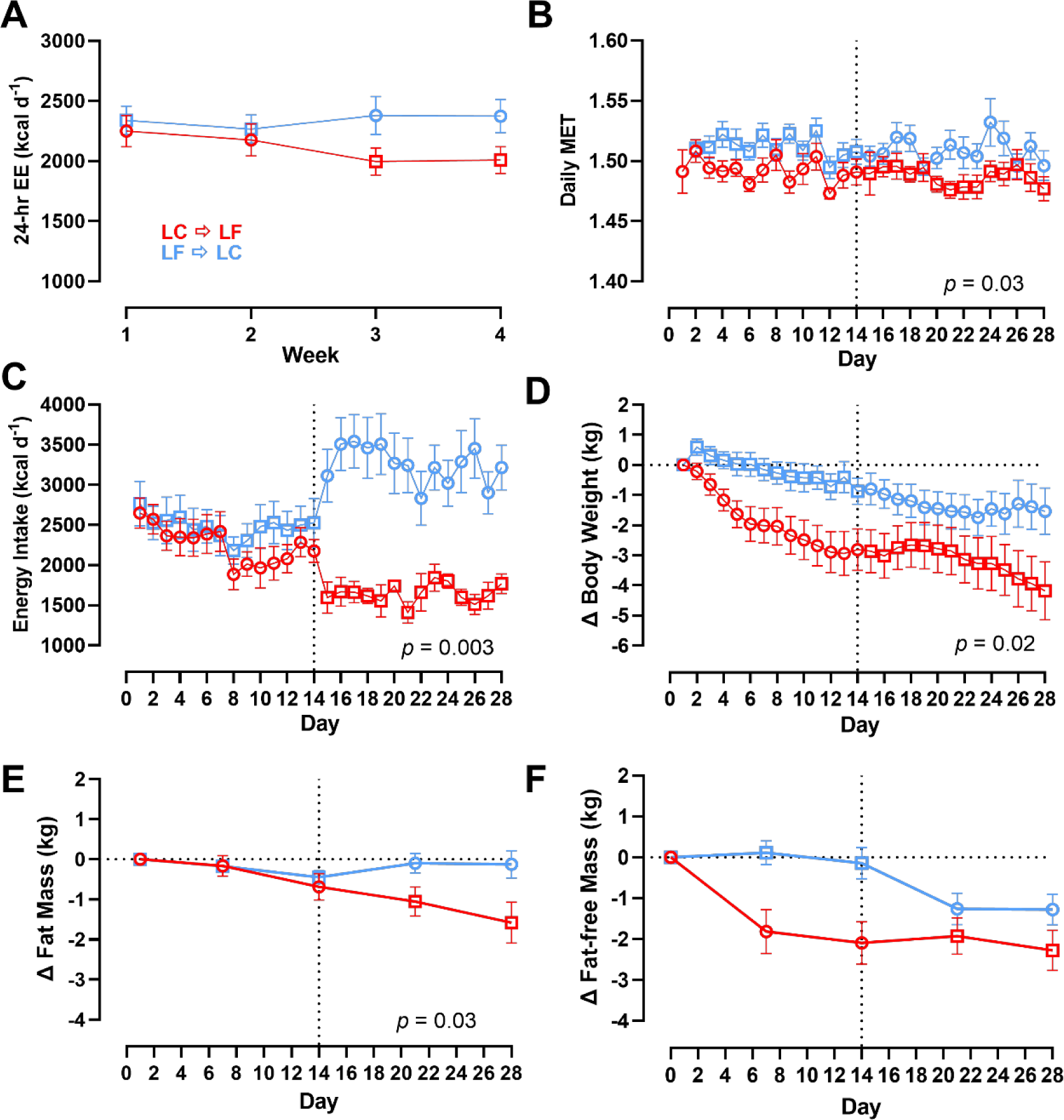
Energy Expenditure, Energy Intake, Body Weight and Composition Changes by Diet Order. Circles represent the LC diet, squares represent the LF diet. Red indicates LC → LF, blue indicates LF → LC. Data are mean ± SEM. *p*-values represent the results of unpaired t-tests. **A)** 24-hr energy expenditure by diet and diet order measured at weeks 1, 2, 3, and 4 over 28-day study period. **B)** Daily MET by diet and diet order over 28-day study period. **C)** Energy intake by diet and diet order over 28-day study period. **D)** Body weight by diet and diet order over 28-day study period. **E)** Fat mass by diet and diet order over 28-day study period. **F)** Fat-f ree mass by diet and diet order over 28-day study period.

#### Energy Intake

As previously reported, there was no significant order effect on within subject differences in mean daily *ad libitum* energy intake between the 2-week LC vs. LF diet periods (143 ± 140 kcal/d; *p* = 0.32) [5]. However, subjects randomized to LC → LF group consumed 922 ± 304 kcal/d less than subjects randomized to LF à LC group over the 28-day study period (*p* = 0.003) (**Figure 2C**). Interestingly, there were no significant diet order group differences in energy intake during the first two weeks (232 ± 306 kcal/d; *p* = 0.44), but the LC → LF group consumed 1610 ± 306 kcal/d less than the LF à LC group over the final 2 weeks (*p* < 0.0001) (**Figure 2C**).

#### Body Weight and Composition

Body weight data are presented in **Figure 2D**. Participants in the LC → LF group lost a total of 4.3 ± 0.8 kg, which was significantly more than the 1.4 ± 0.8 kg weight loss in those who received the LF → LC (*p* = 0.02). Rates of body weight change were calculated during the final week of each diet to allow for equilibration of body fluids over the first week of transitioning to the test diets. Participants randomized to the LC → LF group had greater rates of weight change during both the LC diet period (−0.14 ± 0.03 kg/d for LC → LF vs 0.03 ± 0.03 kg/d for LF → LC; *p* = 0.002) and the LF diet period (−0.18 ± 0.03 kg/d for LC → LF vs −0.08 ± 0.03 kg/d for LF → LC; *p* = 0.02).

**Figure 2E** shows that participants randomized to the LC → LF group lost 1.6 ± 0.4 kg of body fat at the end of the study as compared to only 0.1 ± 0.4 kg of body fat loss for those in the LF → LC group (*p* = 0.03). The LC → LF group lost 2.3 ± 0.4 kg of fat-free mass compared to 1.3 ± 0.4 kg of fat-free mass in the LF → LC group, although this finding was not statistically significant (*p* = 0.12) (**Figure 2F**).

#### Postprandial Responses and Subjective Appetite Measures

With the exception of FFA that tended to be higher in the LC → LF group (p=0.05), circulating postprandial metabolites measured in response to diet-specific meal tests at the end of each LC or LF diet period were not significantly different between the diet order groups (*p* > 0.2; **Supplementary [Figure S1]**). Postprandial active GLP-1 (4.57 ± 0.58 pg·mL^−1^ with LC → LF vs. 4.32 ± 0.58 pg·mL^−1^ with LF → LC; *p* = 0.76), active ghrelin (179.82 ± 20.83 pg·mL^−1^ with LC → LF vs. 143.11 ± 20.83 pg·mL^−1^ with LF → LC; *p* = 0.23), total ghrelin (217.78 ± 37.81 pg·mL^−1^ with LC → LF vs. 227.16 ± 37.81 pg·mL^−1^ with LF → LC; *p* = 0.86), GIP (447.51 ± 50.51 pg·mL^−1^ with LC → LF vs. 449.45 ± 50.52 pg·mL^−1^ with LF → LC; *p* = 0.98), PYY (55.27 ± 4.80 pg·mL^−1^ with LC → LF vs. 60.98 ± 4.80 pg·mL^−1^ with LF → LC; *p* = 0.41), and leptin (29,761.09 ± 7105.95 pg·mL^−1^ with LC → LF vs. 32,270.25 ± 7105.95 pg·mL^−1^ with LF → LC; *p* = 0.81) did not differ between LC → LF vs. LF → LC (**Supplementary [Figure S2]**).

There were no significant diet order differences in food familiarity (69.73 ± 4.41 with LC → LF vs. 81.16 ± 4.41 with LF → LC; *p* = 0.07), hunger (36.14 ± 3.35 with LC → LF vs. 36.39 ± 3.32 with LF → LC; *p* = 0.96), satisfaction (55.32 ± 2.58 with LC → LF vs. 54.46 ± 2.53 with LF → LC; *p* = 0.81), fullness (55.74 ± 2.41 with LC → LF vs. 54.22 ± 2.35 with LF → LC; *p* = 0.65), and eating capacity (41.24 ± 3.21 with LC → LF vs. 41.38 ± 3.18 with LF → LC; *p* = 0.98). However, the LF → LC group reported significantly greater overall ratings of food pleasantness (61.94 ± 3.07 with LC → LF vs. 74.56 ± 3.07 with LF → LC; *p* = 0.004), but this effect was not driven by one diet period over the other (mean difference in LC diet between LC → LF and LF → LC: −9.69 ± 4.87; mean difference in LF diet between LC → LF and LF → LC: −15.51 ± 4.86; *p* = 0.19) (**Supplementary [Figure S3]**).

Meal eating rate data are presented in **Supplementary [Figure S4]**, measured in g/min, was different between the diet order groups during the first 14 days (mean difference: 21.16 ± 6.46 g/min; *p* = 0.001) but similar during the last 14 days (mean difference: 1.46 ± 6.46 g/min; *p* = 0.82). However, over the 28-day period eating rate measured in g/min did not significantly differ by diet order (25.32 ± 4.50 g/min with LC → LF vs. 36.64 ± 4.50 g/min with LF → LC; *p* = 0.08). When measured in kcal/min, eating was faster in LF → LC compared to LC → LF groups (32.49 ± 5.30 kcal/min with LC → LF vs. 47.44 ± 5.30 kcal/min with LF → LC; *p* = 0.046); the observed difference in eating rate in kcal/min occurred mainly in the last two weeks (mean difference: 29.21 ± 7.58 kcal/min; *p* = 0.0001) due to differences in the energy density of the food consumed during that period. These results were not changed when beverages were excluded from the analyses (**Supplementary [Figure S5]**).

**Figure 3A** shows that the energy density of the food consumed did not differ by diet order (LC → LF: 1.41 ± 0.03 kcal/g vs. LF → LC: 1.45 ± 0.03 kcal/g; *p* = 0.40). Overall mass of the food consumed was lower in the LC → LF group (1503 ± 130 g/d) as compared to the LF → LC group (2108 ± 130 g/d; *p* = 0.001) and this was driven by differences during the first 2 weeks (mean difference: 1295 ± 209 g/d; *p* < 0.00001) but not the last 2 weeks (mean difference: 36 ± 209 g/d; *p* = 0.86; **Figure 3B**). **Figure 3C** shows that fiber intake was lower in the LC → LF group (32 ± 4 g/d) as compared to the LF → LC group (49 ± 4 g/d; *p* = 0.0005) over the entire 28 day study period, with differences between the diet order groups being greater during the first 2 weeks (mean difference: 58 ± 5.1 g/d; *p* < 0.00001) than the last 2 weeks (mean difference: 22 ± 5.1 g/d; *p* = 0.00001. These results were not materially changed when beverages were excluded from the analyses (**Supplementary [Figure S6]**).

**Figure 3A-C.**
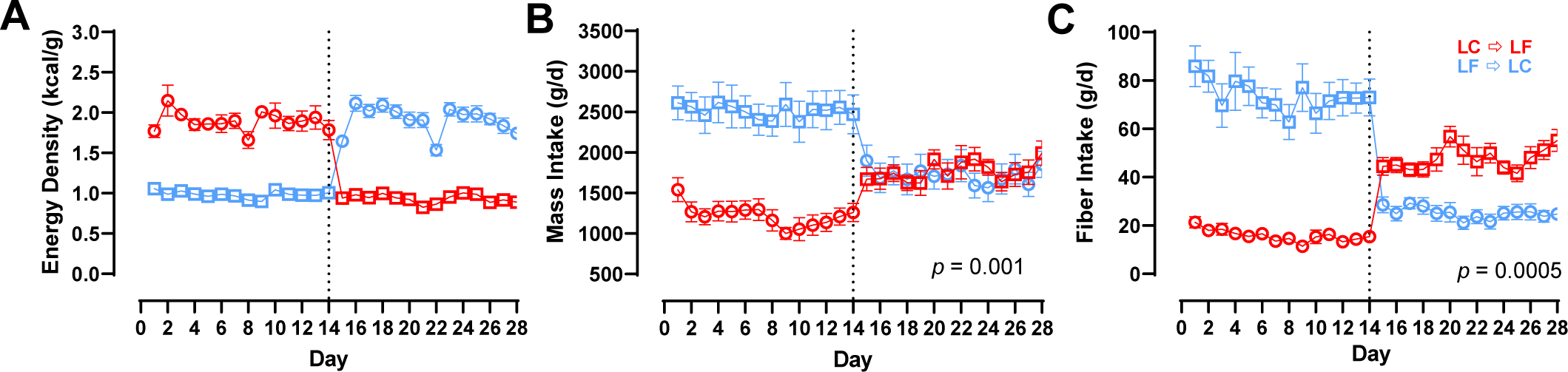
Energy Density, Mass Intake, and Fiber Intake by Diet Order. Circles represent the LC diet, squares represent the LF diet. Red indicates LC → LF, blue indicates LF → LC. Data are mean ± SEM. *p*-values represent the result of unpaired t-tests. **A)** Measured energy density by diet and diet order over 28-day study period. **B)** Mass intake by diet and diet order over 28-day study period. **C)** Fiber intake by diet and diet order over 28-day study period.

#### Ketone and Continuous Glucose Measurements

Capillary β-hydroxybutyrate was significantly greater in the LC → LF group (1.23 ± 0.08 mmol/L) compared to LF → LC group (0.54 ± 0.07 mmol/L; *p* < 0.0001) because the LC diet induced greater concentrations when it was instituted first (**Figure 4A**). Similarly, at the end of the LC diet period plasma total ketones were higher in the LC → LF group (2.34 ± 0.35 mmol/L) compared to LF → LC (1.08 ± 0.33 mmol/L; *p* = 0.01) and 24-hr urine ketones during the LC diet period were higher in the LC → LF group (0.92 ± 0.12 mmol/L) compared to LF → LC (0.52 ± 0.12 mmol/L; *p* = 0.035) (**Figure 4B-C**). Capillary glucose measured via CGM did not differ by diet order (*p* = 0.51; **Supplementary [Figure S7]**).

**Figure 4A-C.**
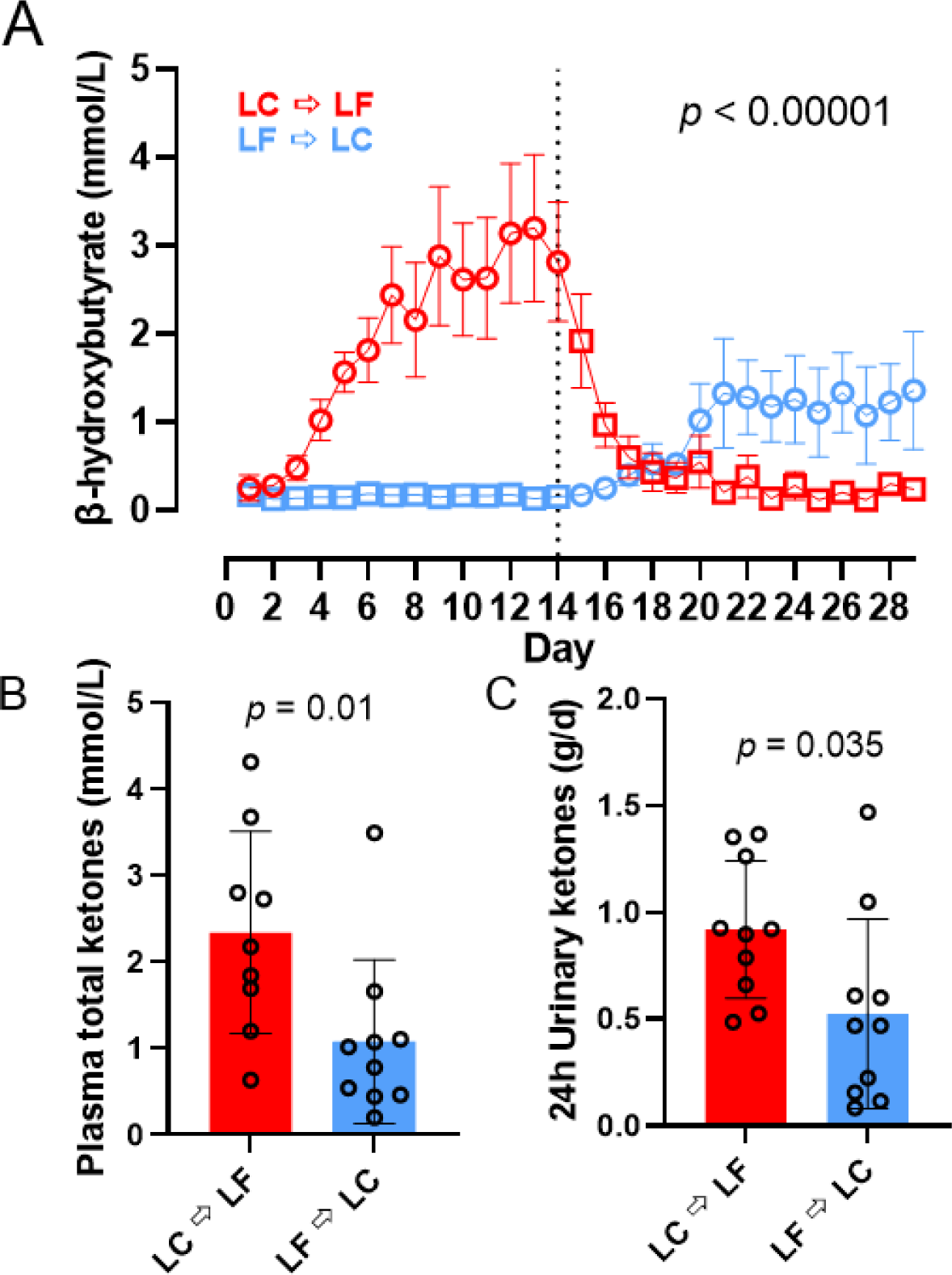
β-hydroxybutyrate, plasma, and urinary ketones. Circles represent the LC diet, squares represent the LF diet. Red indicates LC → LF, blue indicates LF → LC. Data are mean ± SEM. *p*-values represent the result of unpaired t-tests. **A)** β-hydroxybutyrate measured daily in the morning af ter an overnight f ast. **B)** Fasting plasma total ketones measured at the end of each diet period. **C)** 24h urinary ketones measured at the end of each diet period.

During the LC diet, plasma total ketones were negatively correlated with both REE-adjusted energy intake (β = −2.02 mM/kcal; *p* = 0.002) and REE-adjusted CHO intake (β = - 84.23 mM/g; *p* = 0.0009) (**Figure 5A-D**). During the last week of the LC diet, plasma total ketones were negatively correlated with both REE-adjusted energy intake (β = −2.25 mM/kcal/d; *p* = 0.001) and REE-adjusted CHO intake (β = −95.19 mM/g/d; *p* = 0.0003). Neither the slopes nor intercepts differed by diet order (*p* > 0.29). Similar analyses comparing slopes and intercepts for other ketone measurements indicated that differences in REE-adjusted energy or carbohydrate intake was responsible for diet order differences in capillary β-hydroxybutyrate (*p* > 0.70), plasma β-hydroxybutyrate (*p* > 0.38), plasma acetoacetate (*p* > 0.38), and plasma acetone (*p* > 0.25) (**Supplementary [Figure S8 and 9]**).

**Figure 5A-D.**
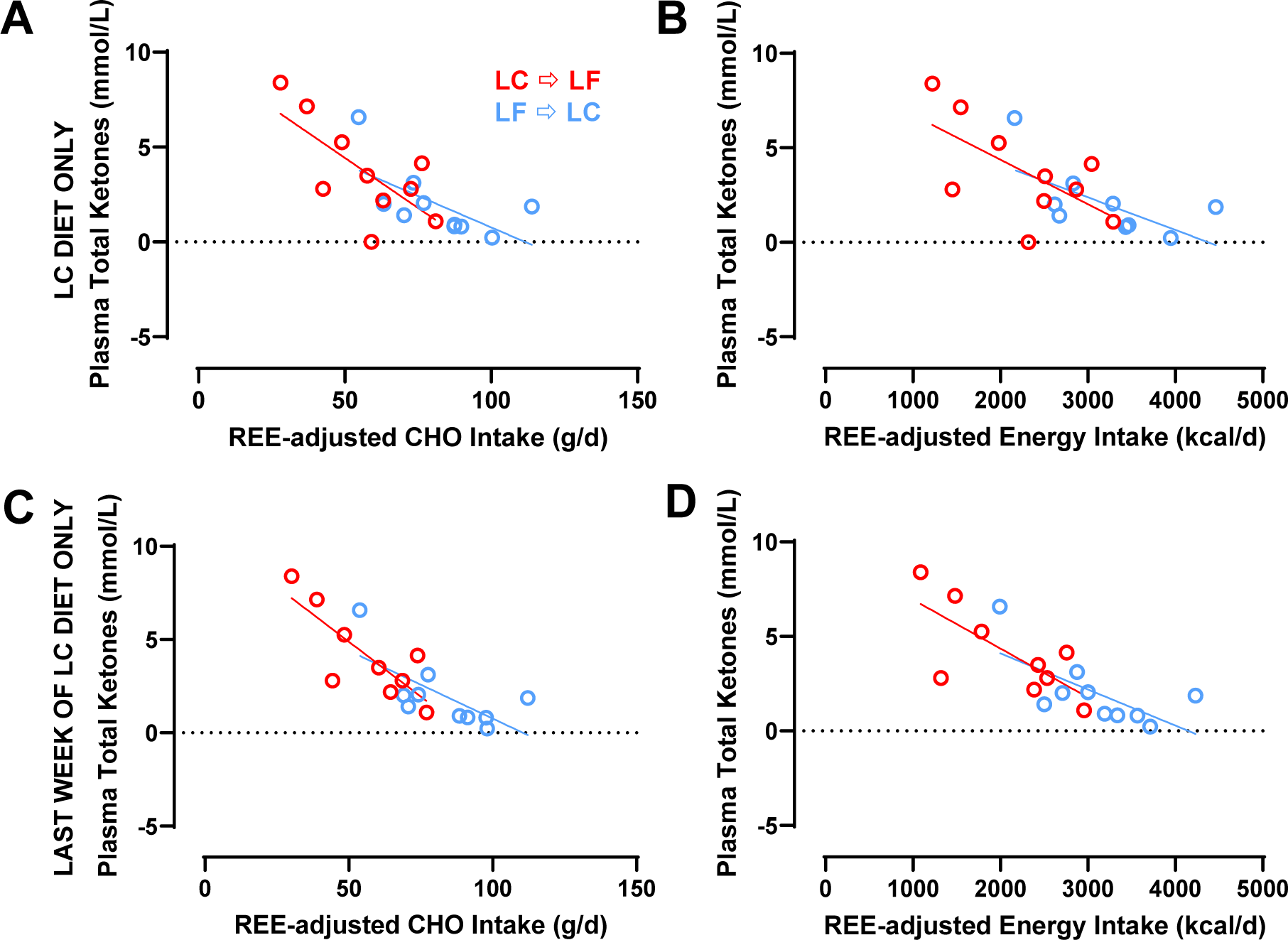
The Relationship Between Plasma Total Ketones and Energy and CHO Intake During the LC Diet Only and During the Last Week of the LC Diet Only. Circles represent the LC diet. Red indicates LC → LF, blue indicates LF → LC. Data are means. **A)** Plasma total ketones and REE-adjusted CHO intake during the 2-week LC diet only. **B)** Plasma total ketones and REE-adjusted energy intake during the 2-week LC diet only. **C)** Plasma total ketones and REE-adjusted CHO intake during the last week of the LC diet only. **D)** Plasma total ketones and REE-adjusted energy intake during the last week of the LC diet only.

### Comparison between UPF→ MPF versus MPF → UPF diet orders

#### Energy Expenditure, Energy Intake, Body Composition, and Continuous Glucose Measurements

Energy expenditure, energy intake, body weight and composition data are presented in **Figure 6A-F**. 24-hr (*p* = 0.87), sleeping (*p* = 0.90), and physical activity energy expenditure (*p* = 0.78) did not differ by diet order. Likewise, Daily METs were not different by diet order (*p* = 0.94). There was no significant diet order effect over the 28-day study period on *ad libitum* energy intake (*p* = 0.51). Similarly, there was no significant diet order effect over the 28-day study period on total weight change (*p* = 0.94) or the rate of body weight change during the final week of the 2-week diets (*p* = 0.70). There was no significant diet order effect over the 28-day study period on fat mass (*p* = 0.47) or fat-free mass change (*p* = 0.89). Interstitial glucose measured via CGM did not differ by diet order (*p* = 0.38; **Supplementary [Figure S10]**).

**Figure 6A-F.**
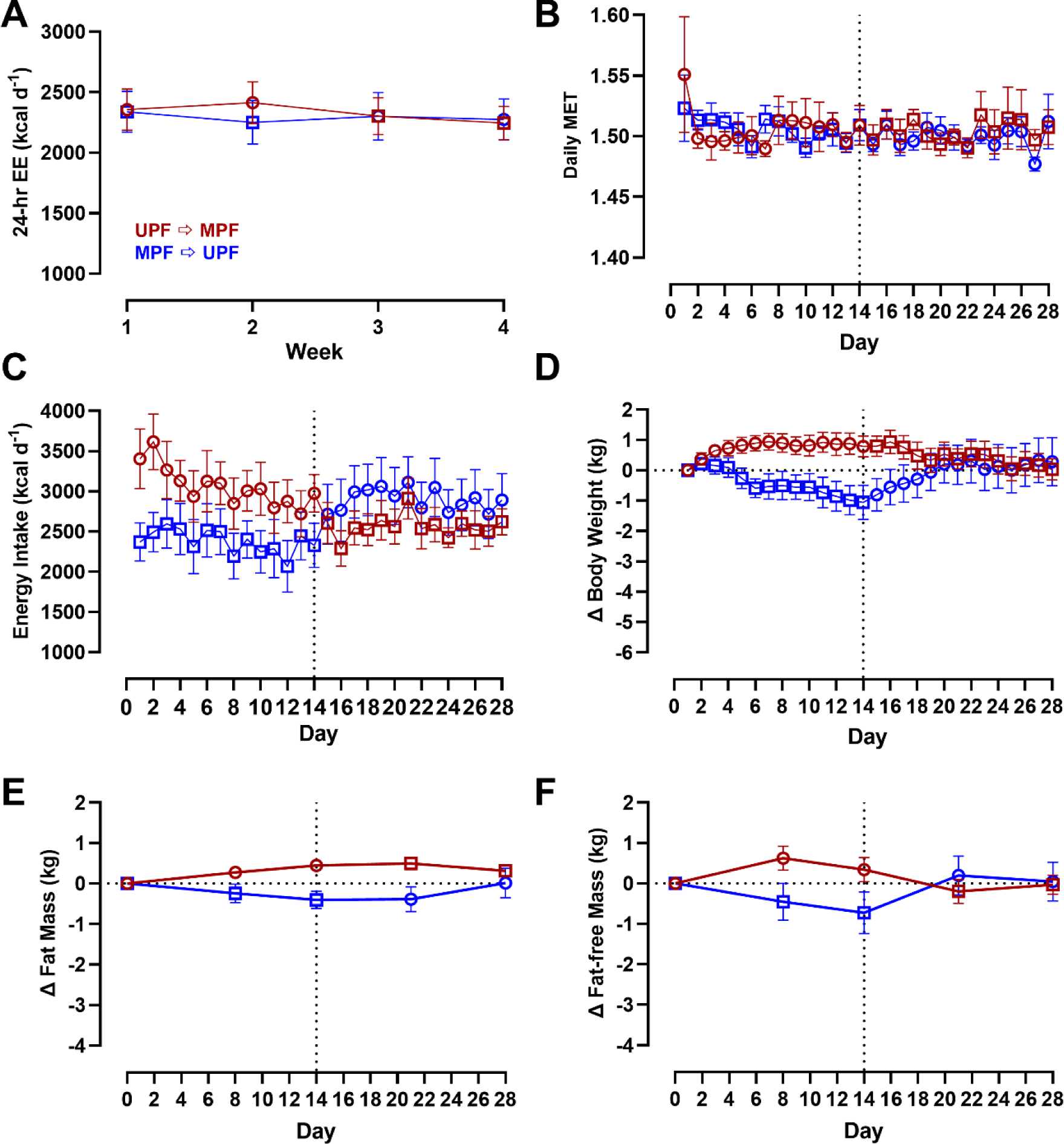
Energy Expenditure, Energy Intake, Body Weight and Composition Changes by Diet Order. Circles represent the UPF diet, squares represent the MPF diet. Red indicates UPF → MPF, blue indicates MPF → UPF. Data are mean ± SEM. Energy intake data are adjusted for resting energy expenditure but presented in figure prior to adjustment. *p*-values represent the results of unpaired t-tests. **A)** 24-hr energy expenditure by diet and diet order measured at weeks 1, 2, 3, and 4 over 28-day study period. **B)** Daily MET by diet and diet order over 28-day study period. **C)** Energy intake by diet and diet order over 28-day study period. **D)** Body weight by diet and diet order over 28-day study period. **E)** Fat mass by diet and diet order over 28-day study period. **F)** Fat-f ree mass by diet and diet order over 28-day study period.

#### Subjective Appetite Measures and Appetite Hormones

There were no significant diet order differences in any measure of subjective appetite, including familiarity (72.36 ± 4.98 with UPF → MPF vs. 72.57 ± 4.96 with MPF → UPF; *p* = 0.98), hunger (36.83 ± 3.89 with UPF → MPF vs. 39.24 ± 3.96 with MPF → UPF; *p* = 0.67), satisfaction (56.82 ± 4.00 with UPF → MPF vs. 55.95 ± 4.07 with MPF → UPF; *p* = 0.88), fullness (56.64 ± 3.99 with UPF → MPF vs. 56.27 ± 4.06 with MPF → UPF; *p* = 0.95), eating capacity (44.76 ± 3.88 with UPF → MPF vs. 46.45 ± 3.93 with MPF → UPF; *p* = 0.76) and pleasantness (64.35 ± 4.22 with UPF → MPF vs. 65.86 ± 4.19 with MPF → UPF; *p* = 0.80) (**Supplementary [Figure S11]**).

Eating rate measured in g/ min (41.13 ± 4.60 g/min with UPF → MPF vs. 32.84 ± 4.60 g/min with MPF → UPF; *p* = 0.20) or kcal/min (50.02 ± 5.31 kcal/min with UPF → MPF vs. 39.70 ± 5.31 kcal/min^−1^ with MPF → UPF; *p* = 0.17) did not differ between diet orders (**Supplementary [Figure S12]**).

Dietary fiber during the UPF period was primarily delivered as a soluble fiber supplement in low calorie beverages served with meals, whereas the dietary fiber during the MPF diet was only in the non-beverage items. Because beverages may act differently than non-beverage foods when it comes to satiety [10], we also analyzed the data excluding beverages. Nevertheless, eating rate with beverages excluded, measured in g/min (*p* = 0.17) or kcal/min (*p* = 0.16), also did not differ by diet order (**Supplementary [Figure S12]**).

Overall energy density did not differ by diet order (UPF → MPF: 1.28 ± 0.06 g/d vs. MPF → UPF: 1.35 ± 0.06 kcal/g; *p* = 0.35), and neither did the mass of food consumed (UPF → MPF: 2263 ± 245 g/d vs. MPF → UPF: 2128 ± 245 g/d; *p* = 0.69) nor fiber consumption (UPF → MPF: 49.2 ± 5.2 g/d vs. MPF → UPF: 45.1 ± 5.2 g/d; *p* = 0.58) differ by diet order (**Figure 7A-C**). Excluding beverages from the analyses, neither the energy density (*p* = 0.12), mass (*p* = 0.66), nor the fiber consumed (*p* = 0.59) resulted in significant diet order differences (**Figure 7D-F**).

**Figure 7A-F.**
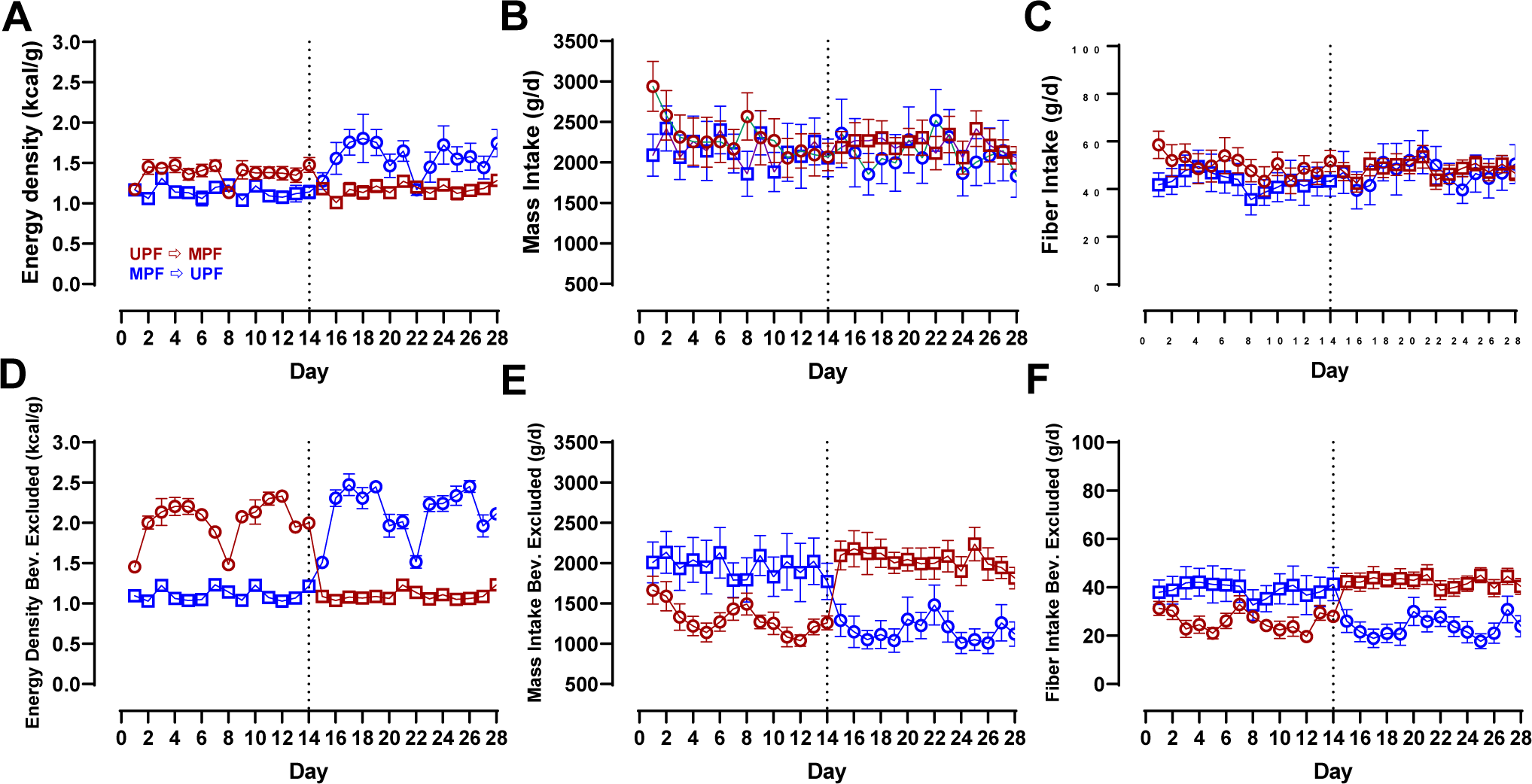
Fiber Intake, Mass Intake, and Energy Density by Diet Order with and without Beverages. Circles represent the UPF diet, squares represent the MPF diet. Red indicates UPF → MPF, blue indicates MPF → UPF. Data are mean ± SEM. *p*-values represent the result of unpaired t-tests. **A)** Measured energy density by diet and diet order over 28-day study period. **B)** Mass intake by diet and diet order over 28-day study period. **C)** Fiber intake by diet and diet order over 28-day study period. **D)** Measured energy density by diet and diet order over 28-day study period with beverages excluded. **E)** Mass intake by diet and diet order over 28-day study period with beverages excluded. **F)** Fiber intake by diet and diet order over 28-day study period with beverages excluded.

## DISCUSSION

The amount of daily food consumed is determined by a wide variety of factors, including properties of the foods available, circulating hormones, metabolic and nutrient needs, as well as a variety of other internal and external signals. Participants in our studies were instructed to eat as much or as little as they desired and they consumed vastly different amounts of energy between LC vs. LF and UPF vs. MPF diets to achieve the same levels of subjective huger, fullness, satisfaction, or eating capacity. While the primary endpoints of the original studies detected no diet order effects on within-subject differences in mean daily *ad libitum* energy intake [4, 5], and there were no significant between diet order group differences in the UPF vs. MPF study, we found substantial differences in energy balance between diet order groups in the LC vs. LF study. Specifically, subjects randomized to the LC → LF group consumed ∼900 fewer kilocalories per day and lost more weight and body fat compared to the LF → LC group, with most of the group differences in energy balance occurring in the last half of the 28-day study following the diet switch.

Why did consuming LF → LC cause greater energy intake and less weight and body fat loss than consuming the reverse LC → LF diet order? One possibility was that the LF → LC diet group found the meals significantly more pleasant on both diets than the LC → LF diet group. However, this explanation is unlikely because the differences in the subjective pleasantness ratings between the diet order groups was exhibited throughout the study whereas differences in food intake occurred between each 14-day study period. Another possibility was that consuming a diet high in carbohydrate content first (the LF diet in the LF → LC group) may have somehow inhibited the development of ketosis during the subsequent LC diet thereby blunting the effect of ketones on appetite suppression [11]. During the LC diet period, ketones in the LF → LC group were significantly lower than in the LC → LF diet group. However, the mathematical relationship between ketones and intake of either energy or carbohydrates was consistent for both diet order groups diet suggesting that differences in ketones during the LC diet were likely driven by differences in food intake between the diet order groups rather than vice versa.

We believe that the most likely explanation of the diet order effects on energy intake involves gut adaptations during the first 14 days, resulting from different intake of fiber and overall mass of food, that carried over to the last 14 days to result in greater overall energy intake in the LF → LC vs the LC → LF group. In other words, LF → LC diet group consumed a large mass of high fiber food with low energy density during the first half of the study that required their gastrointestinal tract to adapt to accommodate a larger mass of high fiber food with low energy density as compared to the LC → LF group whose guts adapted to a smaller mass of low fiber food with high energy density. Energy intake and expenditure was similar between the diet order groups and they lost a similar amount of body fat during those initial 14 days. However, after switching diets, the LF → LC group experienced a diet higher in energy density and lower in fiber, which required greater energy intake to maintain the same subjective appetite as they experienced in the first 14 days. Conversely, the LC → LF group may have adapted to the low mass of food consumed in the first 14 days and when faced with a high fiber diet low in energy density after 14 days a similar mass of food (but lower energy intake) was required to maintain the same subjective appetite. Interestingly, the mass of food consumed by the LF → LC group decreased in the last 14 days, but not to the same low level as in the LC → LF group in the first 14 days despite eating the same diet. Similarly, the mass of the food consumed by the LC → LF group increased during the final 14 days but did not reach the level of the LF → LC group during the first 14 days. Thus, energy intake during the final 14 days was substantially different between the diet order groups despite a similar mass of food consumed between the groups.

It may be that the gut adaptations potentially underlying the order effects were mainly mechanical in nature (e.g., distension of the GI tract), and independent of gut-derived hormones believed to influence appetite because we detected no significant diet order effects on these hormones. Adaptations of the gut microbiome is another possible explanation and such a diet order effect was recently documented in a crossover feeding trial investigating the effect of consuming plant-based meat compared to animal-based meat on serum trimethylamine-N-oxide (TMAO) [12]. While these gut adaptations to LC vs LF diets were rapidly initiated at the start of the study, reversal of their effects was apparently incomplete by the end of the 28-day study suggesting that the time scale of gut adaptations is greater than 14 days.

The lack of significant order effects between the MPF → UPF vs UPF → MPF diet order groups may have been because both diets resulted in a similar mass of food and beverages consumed with similar amounts of fiber. Interestingly, when beverages were excluded from the analyses, the MPF diet had substantially higher fiber, lower energy density, and resulted in a greater mass of non-beverage items consumed as compared to the UPF diet, but there were still no significant differences between the MPF → UPF vs UPF → MPF diet orders. Perhaps this is because beverages consumed with meals can exert important effects on feelings of fullness [13]. Alternatively, perhaps differences in the mass and fiber consumed during the first 14 days need to exceed some threshold to result in differences in gut adaptations that have a significant carryover effect. Indeed, participants randomized to the LF → LC group consumed substantially more fiber and mass of food during the first 14 days on the LF diet than participants in the MPF → UPF group consuming the MPF diet. Similarly, the first 14 days consuming the LC diet in the LC → LF group resulted in somewhat lower fiber and mass intake than the subjects consuming the UPF diet in the UPF → MPF group, even after excluding beverages. Therefore, differences in the mass and fiber intake during the first 14 days were greater between the LF → LC vs LC → LF diet groups than between the MPF → UPF vs UPF → MPF groups regardless of the inclusion or exclusion of beverages. Such differences between the studies may explain why one exhibited a significant effect of diet order whereas the other did not.

The main limitation of the present study is that it was a secondary analysis of data collected for two previous inpatient studies and thus was only exploratory in nature. The original inpatient studies were not designed to investigate order effects between randomized groups or to elucidate the mechanisms. Lastly, while the inpatient nature of the study is a strength, it also limits the generalizability of our results to real-world settings.

Overall, substantial diet order effects on energy balance were observed in the LC vs. LF study but not in the UPF vs. MPF study were likely due to large differences in the mass of food consumed in the first two weeks, related to vastly differing fiber content and energy density between the LC vs. LF diets, that led to gut adaptations carrying over during the subsequent two weeks thereby requiring substantial differences in food intake to maintain the same levels of subjective appetite. Future work should prospectively test this hypothesis, replicate our findings, and examine whether such order effects persist over a longer study durations.

## Data Availability

Data from consenting subjects described in the manuscript, code book, and analytic code will be made publicly and freely available without restriction at: https://osf.io/fjykq/ and https://osf.io/rx6vm/

## Author Contributions

KDH and CMS designed the secondary analysis. JG and CMS analyzed the data. KDH and CMS wrote the manuscript. All authors critically revised the draft and approved the final manuscript.

## SUPPLEMENTAL MATERIALS

**Supplemental Data Figure 1A-F.**
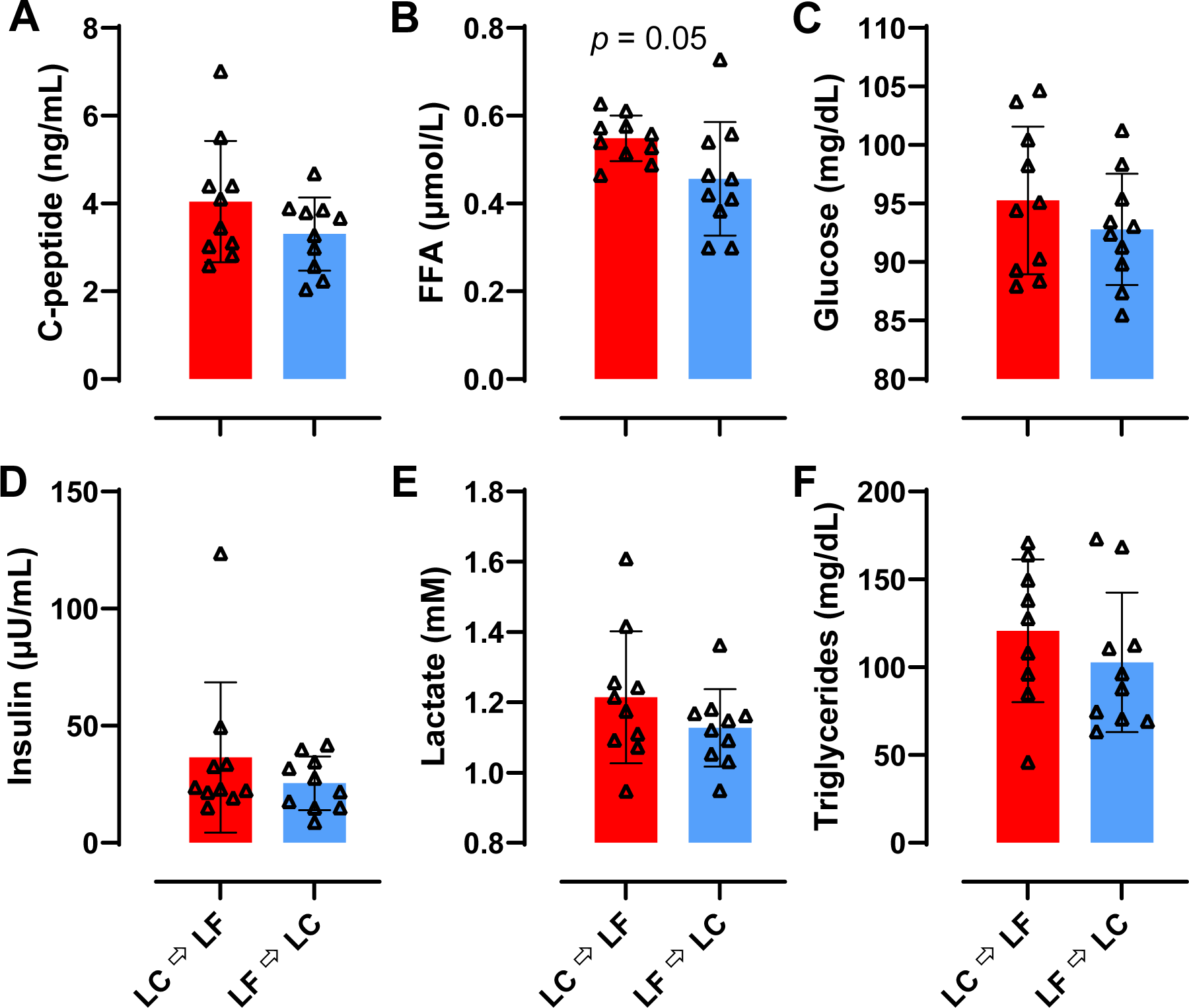
Diet Order Differences in Postprandial Metabolites During LC vs. LF. Red indicates LC → LF, blue indicates LF → LC. Data were collected during mixed meal tests at 0, 10, 20, 30, 60, 90, 120, 180, 240, 300, and 360 minutes post-meal. Data are total area under the curve (tAUC) and reported as mean ± SEM. *p*-values represent the results of unpaired t-tests. **A)** C-peptide tAUC during a mixed meal test. **B)** FFA tAUC during a mixed meal test. **C)** Glucose tAUC during a mixed meal test. **D)** Insulin tAUC during a mixed meal test. **E)** Lactate tAUC during a mixed meal test. **F)** Triglycerides tAUC during a mixed meal test.

**Supplemental Data Figure 2A-F.**
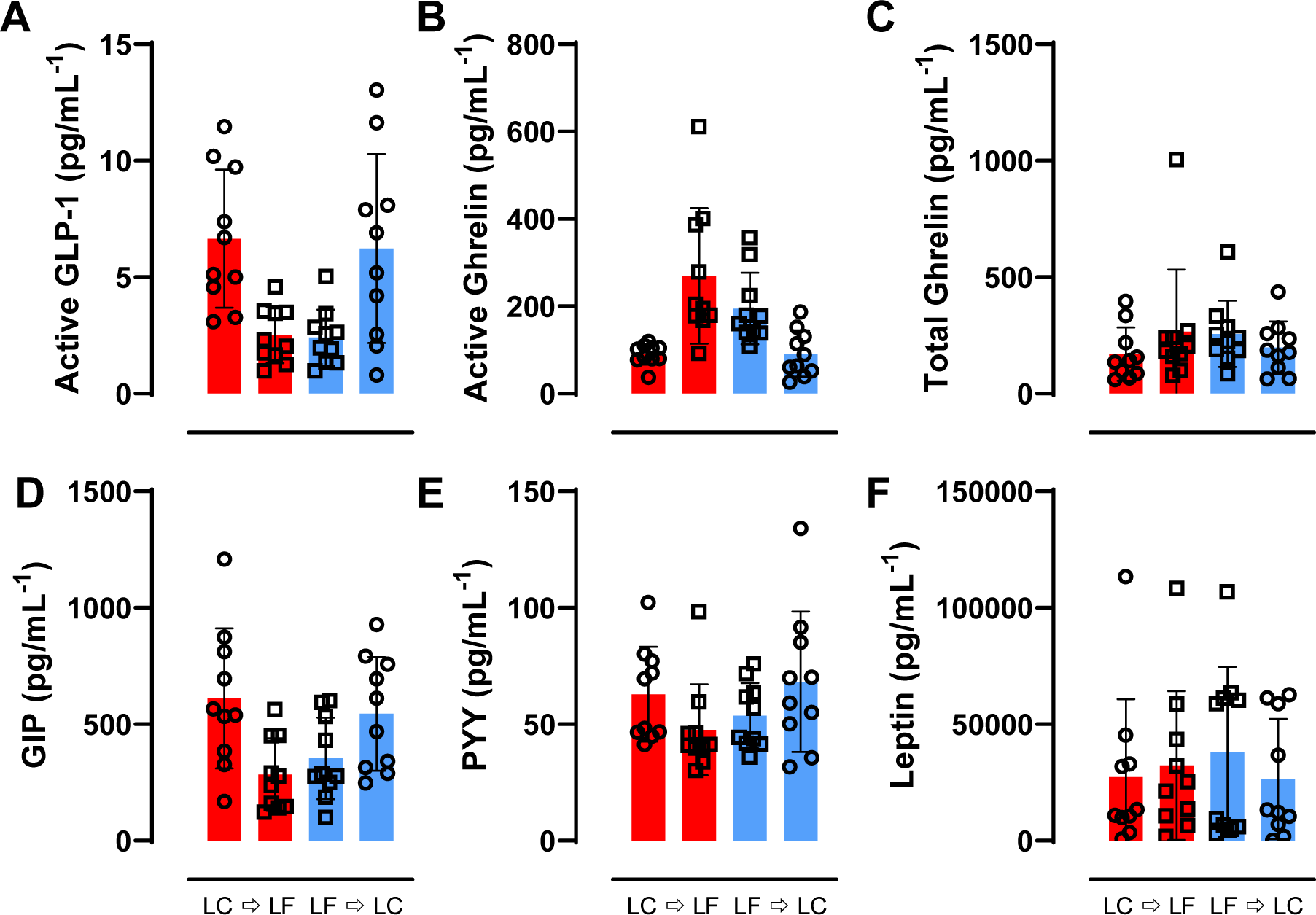
Diet Order Differences in Gut-derived Hormones During a Meal Test During LC vs. LF. Circles represent the LC diet, squares represent the LF diet. Red indicates LC → LF, blue indicates LF → LC. Data were collected during mixed meal tests at 0, 10, 20, 30, 60, 90, 120, 180, 240, 300, and 360 minutes post-meal. Data are total area under the curve (tAUC) and reported as mean ± SEM. *p*-values represent the results of unpaired t-tests. **A)** Active GLP-1 tAUC during a mixed meal test. **B)** Active ghrelin tAUC during a mixed meal test. **C)** Total ghrelin tAUC during a mixed meal test. **D)** GIP tAUC during a mixed meal test. **E)** PYY tAUC during a mixed meal test. **F)** Leptin tAUC during a mixed meal test.

**Supplemental Data Figure 3A-F.**
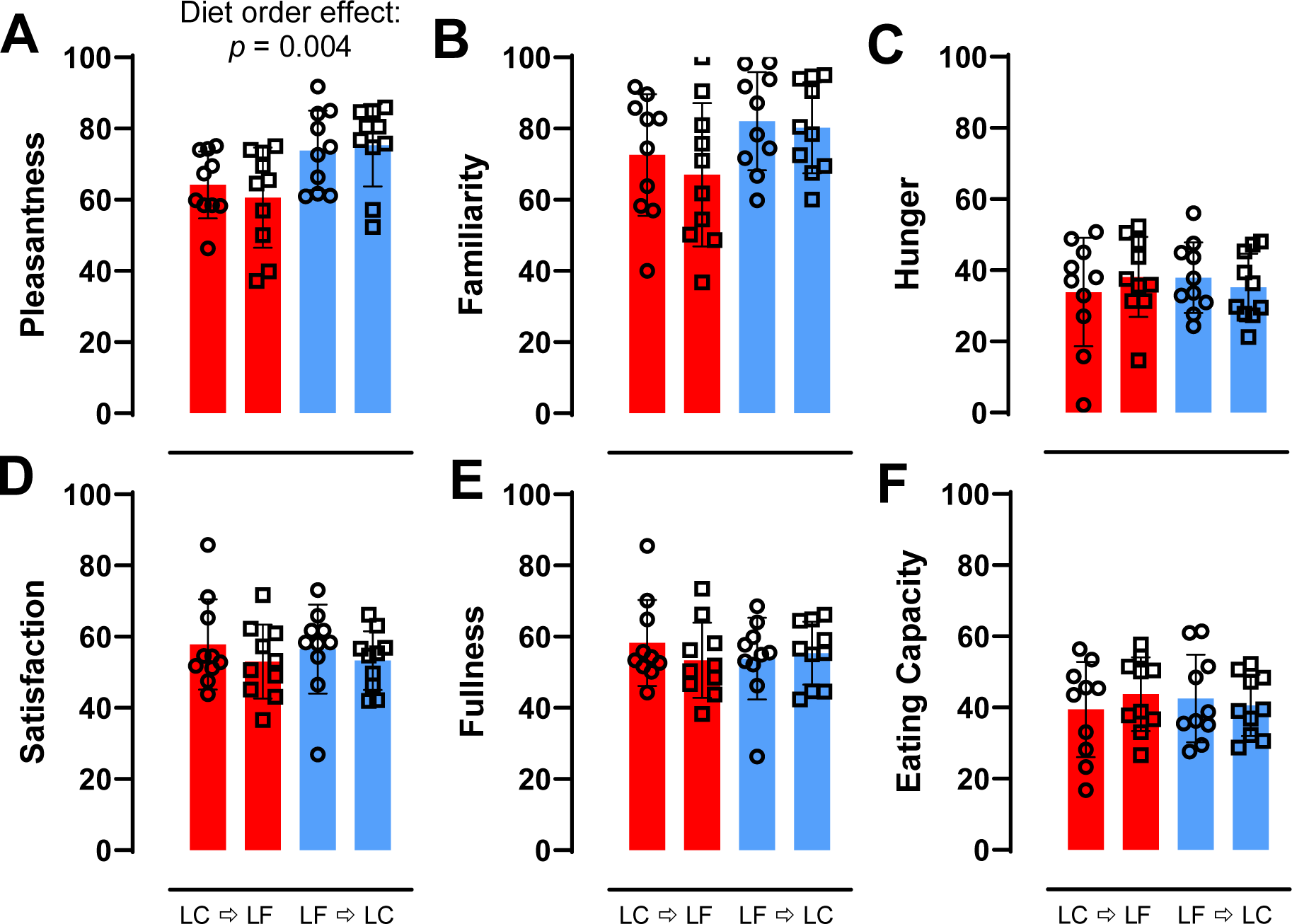
Diet Order Differences in Subjective Measures of Appetite During LC vs. LF. Circles represent the LC diet, squares represent the LF diet. Red indicates LC → LF, blue indicates LF → LC. Data are mean ± SEM. *p*-values represent the results of unpaired t-tests. **A)** Subjective pleasantness of meals by diet order averaged over the 28-day study period. **B)** Subjective familiarity of meals by diet order averaged over the 28-day study period. **C)** Subjective hunger by diet order averaged over the 28-day study period. **D)** Subjective satisfaction by diet order averaged over the 28-day study period. **E)** Subjective fullness by diet order averaged over the 28-day study period. **F)** Subjective eating capacity by diet order averaged over the 28-day study period.

**Supplemental Data Figure 4A-B.**
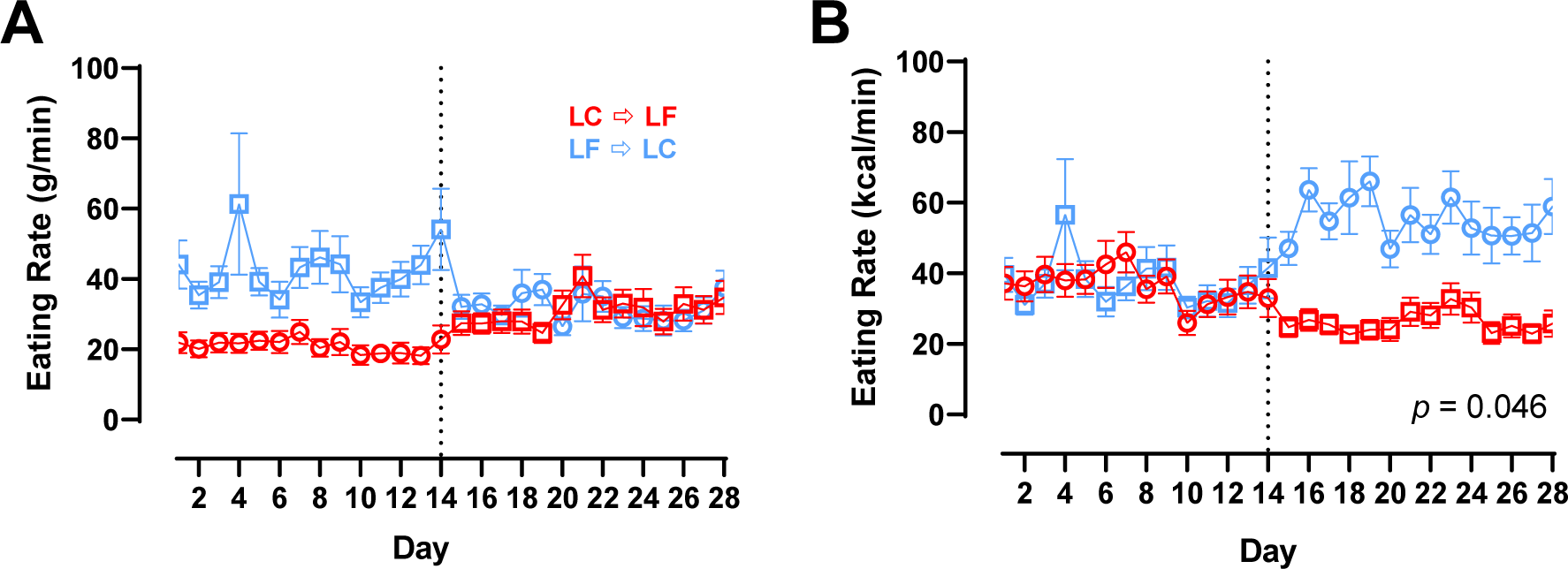
Eating Rate by Diet Order During LC vs. LF. Circles represent the LC diet, squares represent the LF diet. Red indicates LC → LF, blue indicates LF → LC. Data are mean ± SEM. *p*-values represent the result of unpaired t-tests. **A)** Eating rate measured via kcal/min by diet and diet order over 28-day study period. **B)** Eating rate measured via g/min by diet and diet order over 28-day study period.

**Supplemental Data Figure 5A-B.**
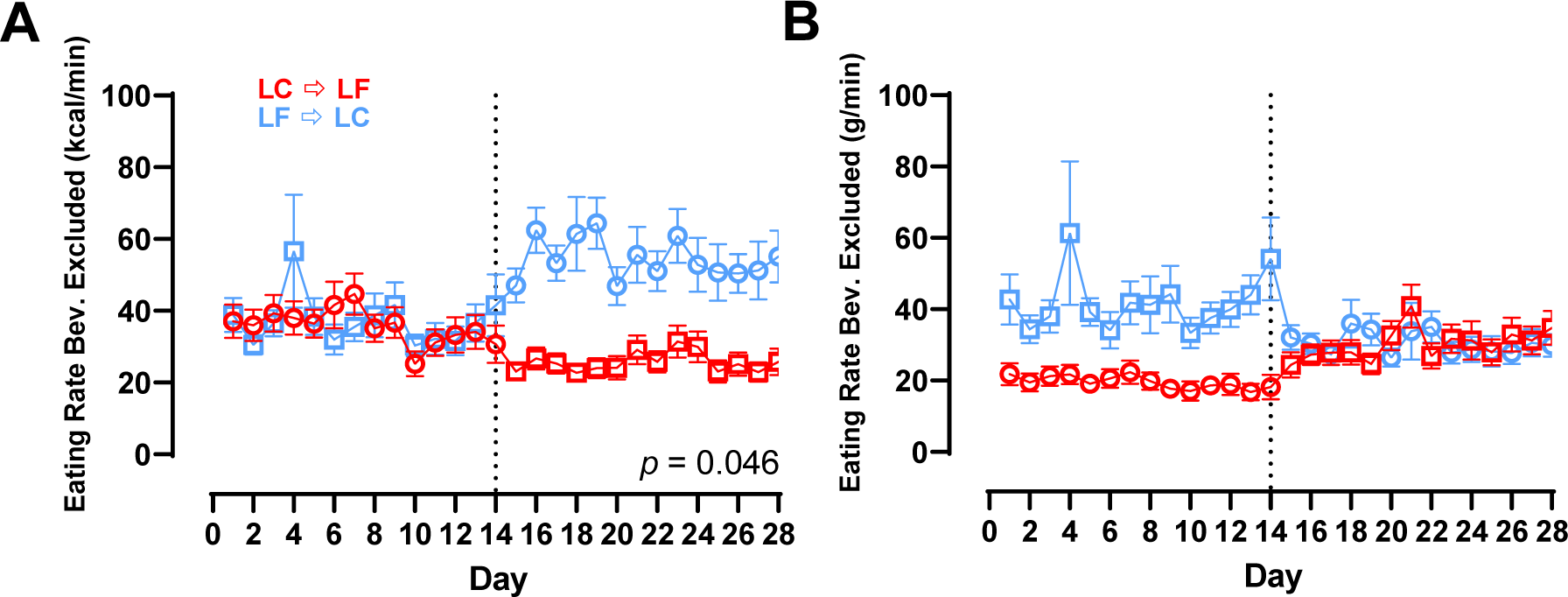
Eating Rate by Diet Order with Beverages Excluded During LC vs. LF. Circles represent the LC diet, squares represent the LF diet. Red indicates LC → LF, blue indicates LF → LC. Data are mean ± SEM. *p*-values represent the result of unpaired t-tests. **A)** Eating rate beverages excluded measured via kcal/min^−1^ by diet and diet order over 28-day study period. **B)** Eating rate beverages excluded measured via g/min^−1^ by diet and diet order over 28-day study period.

**Supplemental Data Figure 6A-C.**
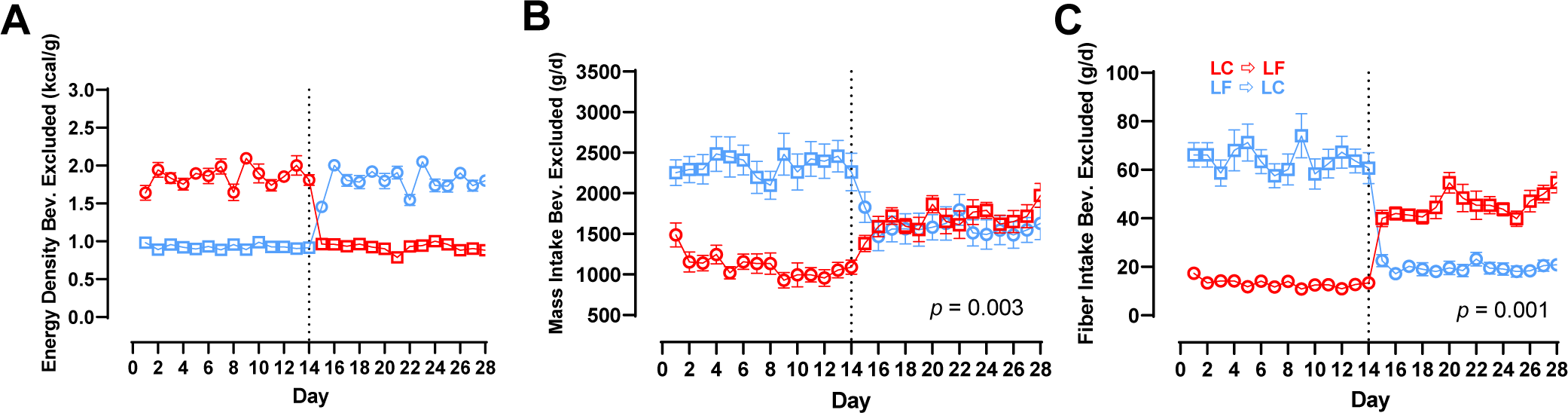
Energy Density, Mass Intake, and Fiber Intake by Diet Order with Beverages Excluded During LC vs. LF. Circles represent the LC diet, squares represent the LF diet. Red indicates LC → LF, blue indicates LF → LC. Data are mean ± SEM. *p*-values represent the result of unpaired t-tests. **A)** Measured energy density beverages excluded by diet and diet order over 28-day study period. **B)** Mass intake beverages excluded by diet and diet order over 28-day study period. **C)** Fiber intake beverages excluded by diet and diet order over 28-day study period.

**Supplemental Data Figure 7.**
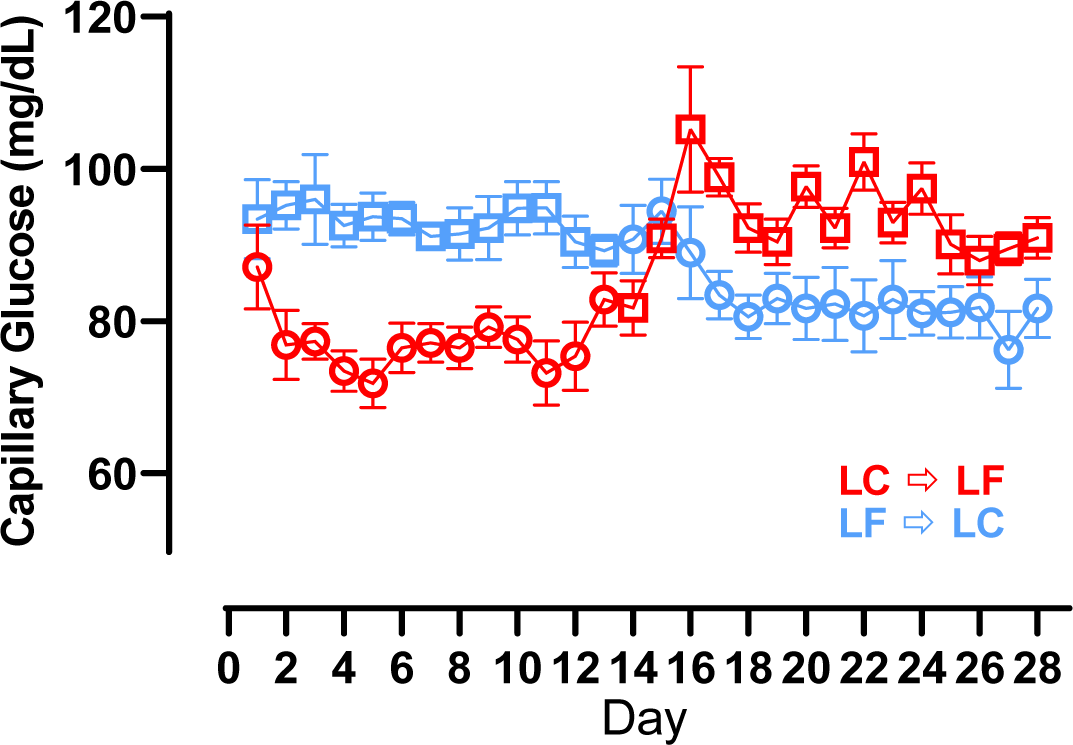
Capillary Glucose Measured via CGM During LC vs. LF. Circles represent the LC diet, squares represent the LF diet. Red indicates LC → LF, blue indicates LF → LC. Data are mean ± SEM. *p*-values represent the results of unpaired *t*-tests.

**Supplemental Data Figure 8A-H.**
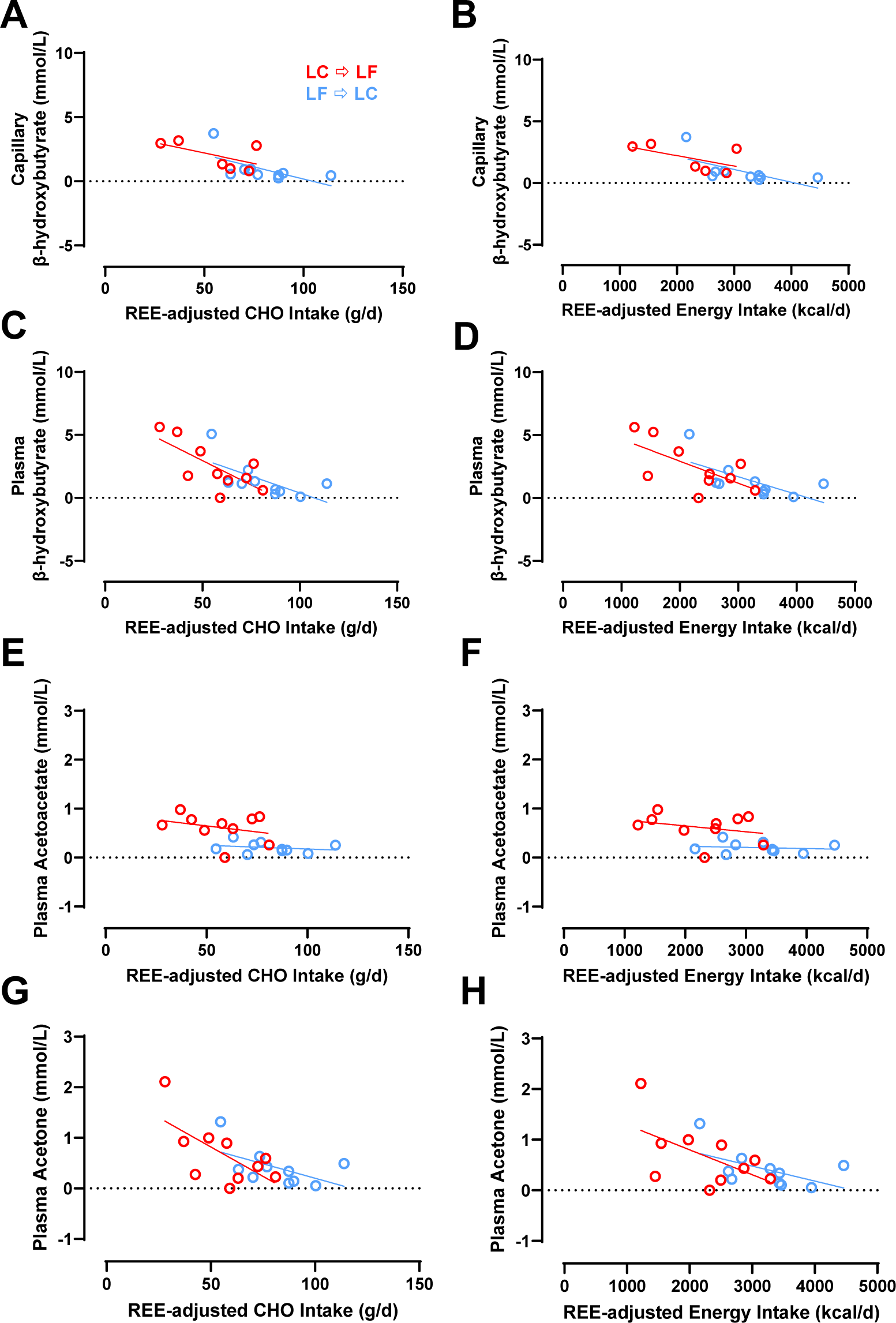
The Relationship Between Capillary β-hydroxybutyrate, Plasma β-hydroxybutyrate, Plasma Acetoacetate, and Plasma Acetone and Energy and CHO Intake During the LC Diet Only. Circles represent the LC diet. Red indicates LC → LF, blue indicates LF → LC. Data are means. **A)** Capillary β-hydroxybutyrate and REE-adjusted CHO intake during the 2-week LC diet only. **B)** Capillary β-hydroxybutyrate and REE-adjusted energy intake during the 2-week LC diet only. **C)** Plasma β-hydroxybutyrate and REE-adjusted CHO intake during the 2-week LC diet only. **D)** Plasma β-hydroxybutyrate and REE-adjusted energy intake during the 2-week LC diet only. **E)** Plasma Acetoacetate and REE-adjusted CHO intake during the 2-week LC diet only. **F)** Plasma Acetoacetate and REE-adjusted energy intake during the 2-week LC diet only. **G)** Plasma Acetone and REE-adjusted CHO intake during the 2-week LC diet only. **H)** Plasma Acetone and REE-adjusted energy intake during the 2-week LC diet only.

**Supplemental Data Figure 9A-H.**
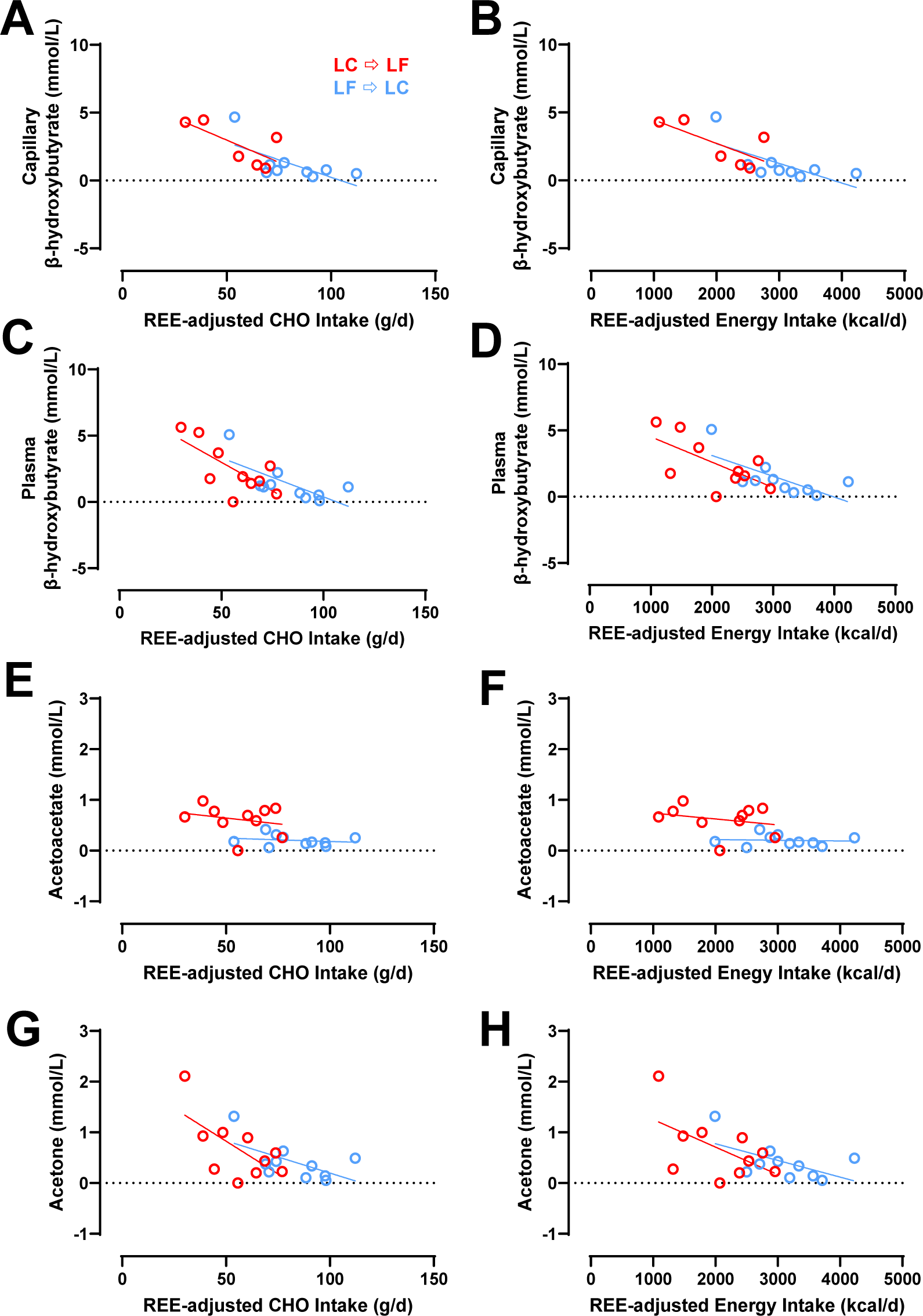
The Relationship Between Capillary β-hydroxybutyrate, Plasma β-hydroxybutyrate, Plasma Acetoacetate, and Plasma Acetone and Energy and CHO Intake During the Last Week of the LC Diet Only. Circles represent the LC diet. Red indicates LC → LF, blue indicates LF → LC. Data are means. **A)** Capillary β-hydroxybutyrate and REE-adjusted CHO intake during the last week of the LC diet only. **B)** Capillary β-hydroxybutyrate and REE-adjusted energy intake during the last week of the LC diet only. **C)** Plasma β-hydroxybutyrate and REE-adjusted CHO intake during the last week of the LC diet only. **D)** Plasma β-hydroxybutyrate and REE-adjusted energy intake during the last week of the LC diet only. **E)** Plasma Acetoacetate and REE-adjusted CHO intake during the last week of the LC diet only. **F)** Plasma Acetoacetate and REE-adjusted energy intake during the last week of the LC diet only. **G)** Plasma Acetone and REE-adjusted CHO intake during the last week of the LC diet only. **H)** Plasma Acetone and REE-adjusted energy intake during the last week of the LC diet only.

**Supplemental Data Figure 10.**
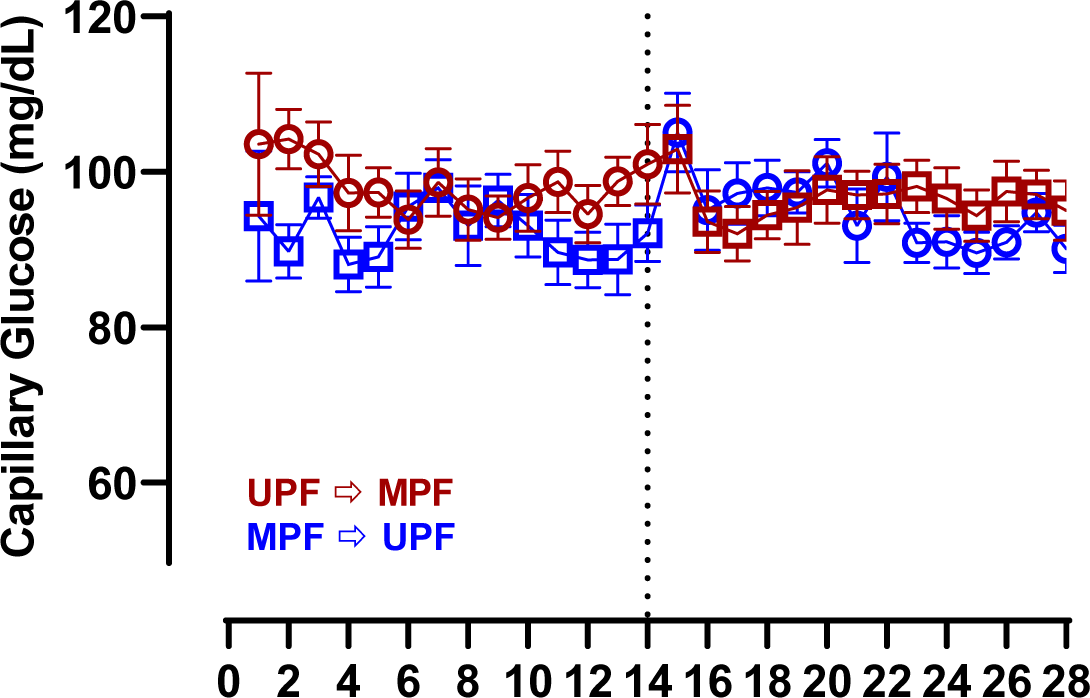
Capillary Glucose Measured via CGM During UPF vs. MPF. Circles represent the UPF diet, squares represent the MPF diet. Red indicates UPF → MPF, blue indicates MPF → UPF. Data are mean ± SEM.

**Supplemental Data Figure 11A-F.**
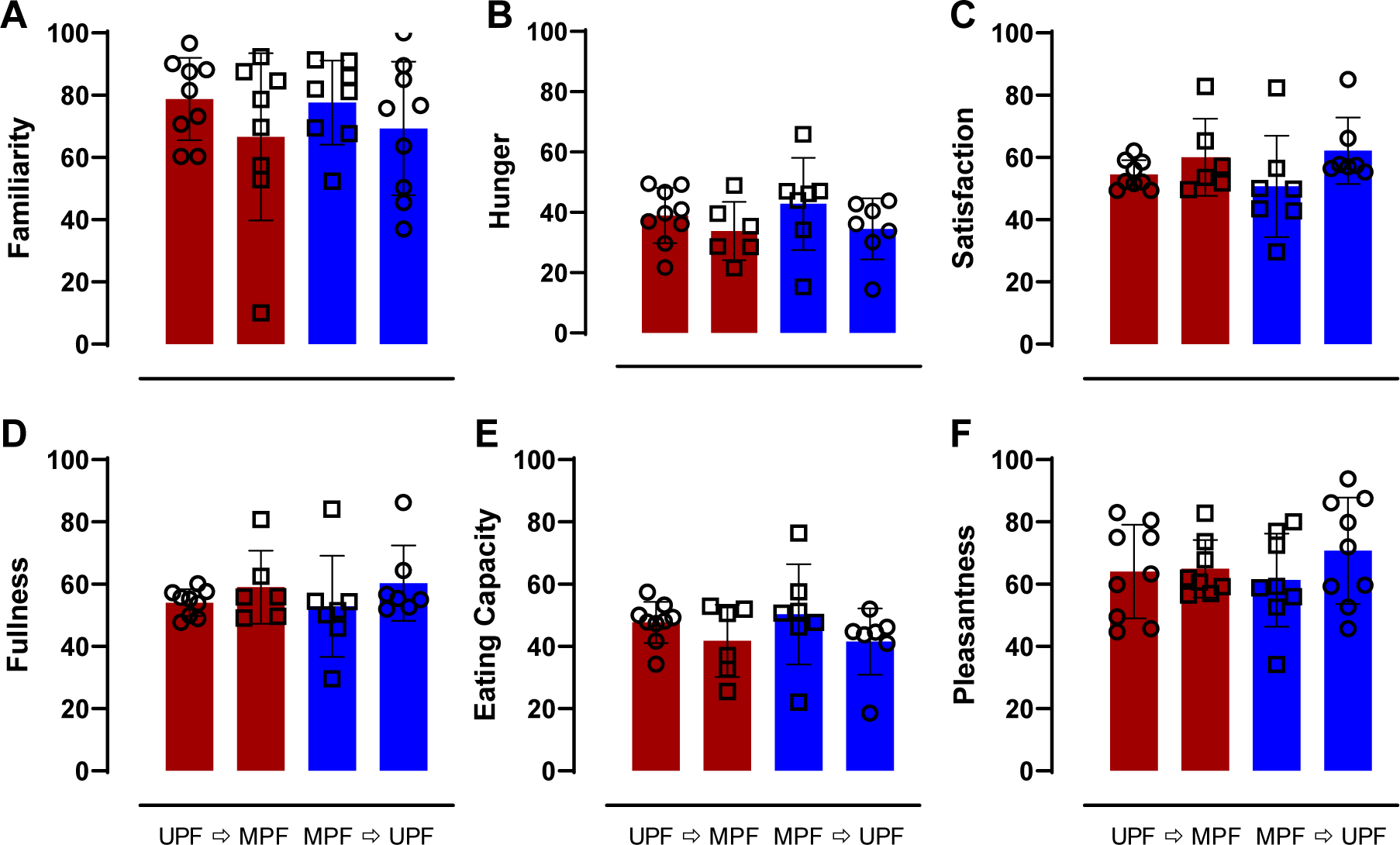
Diet Order Diff erences in Subjective Measures of Appetite During UPF vs. MPF. Circles represent the UPF diet, squares represent the MPF diet. Red indicates UPF → MPF, blue indicates MPF → UPF. Data are mean ± SEM. **A)** Subjective familiarity of meals by diet order averaged over the 28-day study period. *p*-values represent the results of unpaired t-tests. **B)** Subjective hunger by diet order averaged over the 28-day study period. **C)** Subjective satisfaction by diet order averaged over the 28-day study period. **D)** Subjective f ullness by diet order averaged over the 28-day study period. **E)** Subjective eating capacity by diet order averaged over the 28-day study period. **F)** Subjective pleasantness of meals by diet order averaged over the 28-day study period.

**Supplemental Data Figure 12A-D.**
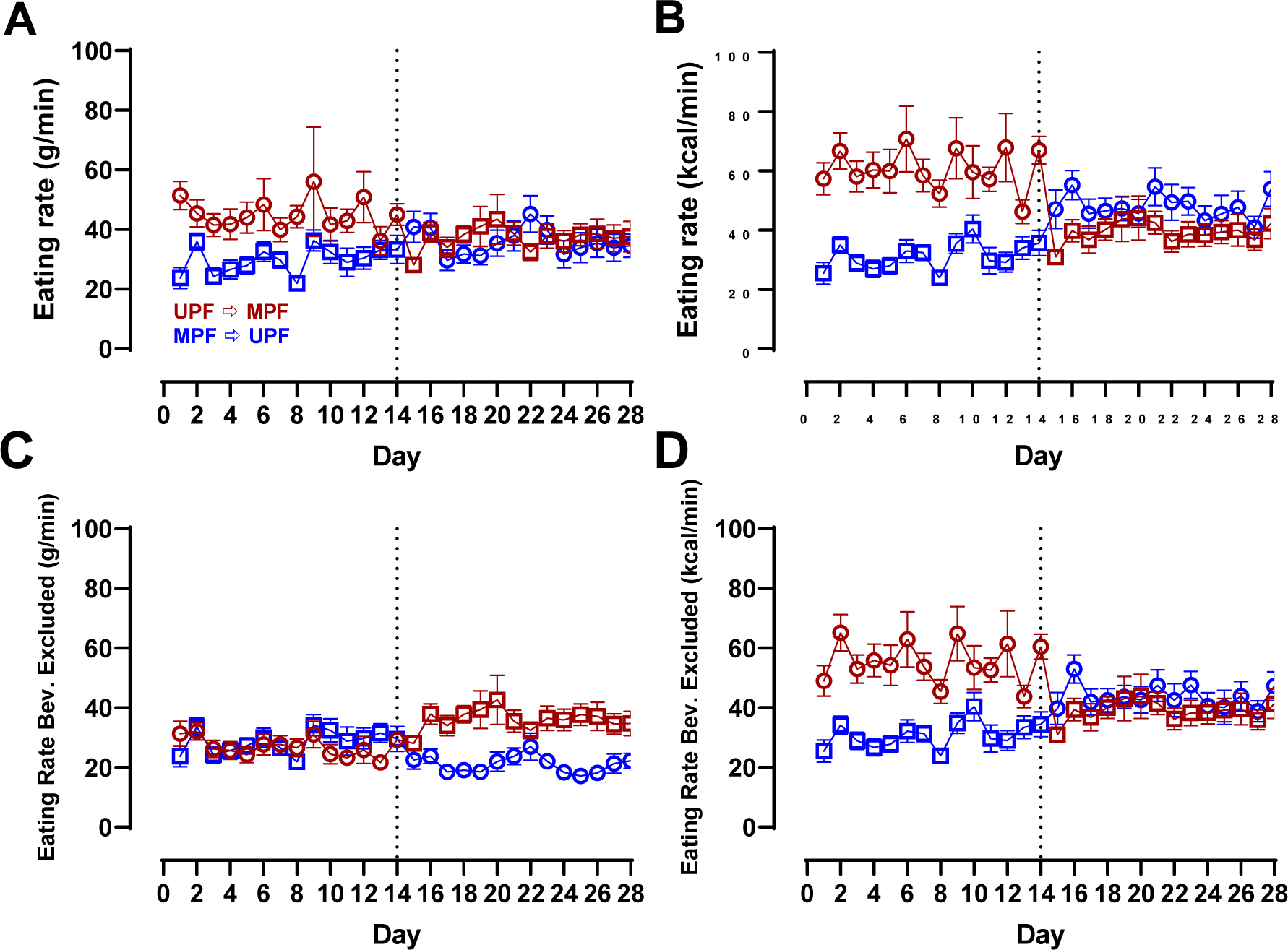
Eating Rate by Diet Order During UPF vs. MPF. Circles represent the UPF diet, squares represent the MPF diet. Red indicates UPF → MPF, blue indicates MPF → UPF. Data are mean ± SEM. **A)** Eating rate measured via g/min by diet and diet order over 28-day study period. **B)** Eating rate measured via kcal/min by diet and diet order over 28-day study period. **C)** Eating rate measured via g/min by diet and diet order over 28-day study period with beverages excluded. **D)** Eating rate measured via kcal/min by diet and diet order over 28-day study period with beverages excluded.

## REFERENCES

1. Lichtenstein AH, Petersen K, Barger K, Hansen KE, Anderson CA, Baer DJ, Lampe JW, Rasmussen H, Matthan NR: Perspective: design and conduct of human nutrition randomized controlled trials. Advances in Nutrition 2021, 12(1):4–20.

2. Hall KD: Challenges of human nutrition research. Science 2020, 367(6484):1298–1300.

3. Dwan K, Li T, Altman DG, Elbourne D: CONSORT 2010 statement: extension to randomised crossover trials. BMJ 2019, 366:l4378.

4. Hall KD, Ayuketah A, Brychta R, Cai H, Cassimatis T, Chen KY, Chung ST, Costa E, Courville A, Darcey V: Ultra-processed diets cause excess calorie intake and weight gain: an inpatient randomized controlled trial of ad libitum food intake. Cell metabolism 2019, 30(1):67–77. e63.

5. Hall KD, Guo J, Courville AB, Boring J, Brychta R, Chen KY, Darcey V, Forde CG, Gharib AM, Gallagher I: Effect of a plant-based, low-fat diet versus an animal-based, ketogenic diet on ad libitum energy intake. Nature medicine 2021, 27(2):344–353.

6. Monteiro CA, Cannon G, Moubarac J-C, Levy RB, Louzada MLC, Jaime PC: The UN Decade of Nutrition, the NOVA food classification and the trouble with ultra-processing. Public health nutrition 2018, 21(1):5–17.

7. Flood A, Subar AF, Hull SG, Zimmerman TP, Jenkins DJ, Schatzkin A: Methodology for adding glycemic load values to the National Cancer Institute Diet History Questionnaire database. Journal of the American Dietetic Association 2006, 106(3):393–402.

8. Schoffelen PF, Westerterp KR: Intra-individual variability and adaptation of overnight-and sleeping metabolic rate. Physiology & behavior 2008, 94(2):158–163.

9. Hall KD, Chen KY, Guo J, Lam YY, Leibel RL, Mayer LE, Reitman ML, Rosenbaum M, Smith SR, Walsh BT: Energy expenditure and body composition changes after an isocaloric ketogenic diet in overweight and obese men. The American journal of clinical nutrition 2016, 104(2):324–333.

10. Stribiţcaia E, Evans CE, Gibbons C, Blundell J, Sarkar A: Food texture influences on satiety: systematic review and meta-analysis. Scientific reports 2020, 10(1):12929.

11. Ludwig DS, Aronne LJ, Astrup A, de Cabo R, Cantley LC, Friedman MI, Heymsfield SB, Johnson JD, King JC, Krauss RM: The carbohydrate-insulin model: a physiological perspective on the obesity pandemic. The American journal of clinical nutrition 2021, 114(6):1873–1885.

12. Crimarco A, Springfield S, Petlura C, Streaty T, Cunanan K, Lee J, Fielding-Singh P, Carter MM, Topf MA, Wastyk HC: A randomized crossover trial on the effect of plant-based compared with animal-based meat on trimethylamine-N-oxide and cardiovascular disease risk factors in generally healthy adults: Study With Appetizing Plantfood—Meat Eating Alternative Trial (SWAP-MEAT). The American journal of clinical nutrition 2020, 112(5):1188–1199.

13. DellaValle DM, Roe LS, Rolls BJ: Does the consumption of caloric and non-caloric beverages with a meal affect energy intake? Appetite 2005, 44(2):187–193.

